# Adversarial Validation Reveals Diagnostic Workflow Leakage in PCOS Machine Learning Models

**DOI:** 10.64898/2026.07.22.26358682

**Authors:** Kornelia Kataryńczuk, Aleksandra Stachowiak, Natalia Piórkowska, Alan Ostromęcki, Grzegorz Franik, Anna Bizo

## Abstract

**Background:** Machine-learning models for polycystic ovary syndrome (PCOS) and other conditions frequently report near-perfect diagnostic performance, but retrospective datasets assembled from routine clinical practice can encode diagnostic-group membership in how data were acquired rather than in disease biology, and this acquisition-related information can be indistinguishable from genuine clinical signal under conventional validation.

**Objective:** To determine, using a real-world PCOS cohort as a case study, whether high classification performance reflected clinically meaningful information or artifacts of data provenance, schema structure, and measurement-acquisition workflow, and to develop a generalizable audit framework for detecting such artifacts in retrospective medical machine learning.

**Methods:** We analyzed 1,331 retrospective records (1,286 PCOS, 45 controls) from a single endocrine-gynecology database. A layered acquisition-bias framework compared classification performance using (i) raw and harmonized missingness patterns alone, (ii) measured values with and without explicit missingness indicators, and (iii) ascertainment-balanced feature sets with and without age. Logistic regression and random forest were evaluated using repeated stratified cross-validation, bootstrap resampling, label-permutation testing, and calibration analysis, and the framework was validated against a semi-synthetic experiment with known ground truth.

**Results:** Diagnostic status was perfectly predicted (ROC-AUC = 1.000) from missingness patterns alone, before any clinical value was examined, and this persisted after semantic harmonization of duplicated source columns. Performance declined progressively as acquisition-sensitive information was removed, from near-ceiling in raw and harmonized value models to a mean ROC-AUC of approximately 0.80–0.82 in the most restrictive ascertainment-balanced, age-excluded representation. The semi-synthetic experiment reproduced this pattern under known data-generating conditions, confirming that harmonization removes schema-fragmentation artifacts but not workflow-driven acquisition bias.

**Conclusions:** Apparent diagnostic performance in this cohort was substantially attributable to diagnostic workflow and data-acquisition structure rather than to a stable, transportable biological signal. The layered audit framework generalizes beyond PCOS and offers a practical tool for detecting acquisition-related leakage in retrospective clinical machine-learning studies.

## 1. Introduction

Machine learning is increasingly applied to routinely collected clinical data to support diagnosis and risk stratification, and polycystic ovary syndrome (PCOS) has become a popular case study for this approach. PCOS is a common endocrine-metabolic disorder diagnosed under the Rotterdam ESHRE/ASRM criteria by the presence of at least two of three features - oligo- or anovulation, clinical or biochemical hyperandrogenism, and polycystic ovarian morphology - after exclusion of alternative causes [1]. Because the syndrome is heterogeneous and no single test is diagnostic, a substantial literature has explored whether supervised learning applied to clinical, hormonal, and metabolic variables can support or accelerate diagnosis. A recent systematic review identified dozens of such studies, several reporting classification accuracies above 95% [2], and models trained on public PCOS datasets have reported similarly high performance from routinely collected clinical and hormonal features [3].

Reported accuracies of this magnitude are difficult to reconcile with the acknowledged clinical difficulty of PCOS diagnosis and raise the question of what these models are actually learning. A growing body of evidence from other areas of medical machine learning suggests that very high apparent performance can arise from “shortcut learning”: models exploiting incidental, easy-to-detect statistical regularities in a dataset rather than the intended clinical signal [4]. Convolutional networks trained to detect pneumonia or COVID-19 from chest radiographs, for example, have been shown to instead learn to recognize the hospital system, scanner type, or department that produced the image, because these acquisition-related cues correlated with disease prevalence in the training data [5,6]. Because such shortcuts can produce excellent held-out performance within a single dataset, they are frequently invisible to conventional internal validation and are only revealed when the source of information is deliberately interrogated or an external cohort is tested [7,8].

Retrospective databases assembled from real-world clinical practice, such as the PCOS cohort analyzed here, are particularly susceptible to this failure mode. Because record-level testing decisions reflect clinical workflow rather than a prespecified study protocol, the presence or absence of a given measurement is rarely random: clinicians order tests differently for patients with different working diagnoses, and comparison or control groups drawn from unrelated clinical pathways are frequently characterized by systematically different test panels. This produces “informative missingness”, in which the pattern of which variables were recorded— independent of their measured values - carries predictive information [9,10]. Informative missingness is not inherently a methodological error; when a model is deployed in the same clinical workflow that generated its training data, it can be a legitimate source of predictive power [9]. However, when the objective of a study is to characterize the biological or clinical signal that distinguishes a disease from its comparators, rather than to reproduce the operational behavior of a specific clinic, acquisition-related missingness becomes a source of leakage that can be mistaken for diagnostic accuracy.

A parallel and closely related problem arises from the raw structure of clinical databases. Source systems frequently duplicate the same clinical concept across several differently named columns, reflecting changes in laboratory equipment, reporting conventions, or data-entry practice over time; if such columns are populated more often for one diagnostic group than another, their pattern of availability can itself function as an unintended identifier of group membership, independent of any underlying clinical measurement. Distinguishing this kind of schema-level artifact from clinically meaningful workflow-related missingness requires an audit framework capable of separating the two mechanisms rather than treating “missingness” as a single, undifferentiated phenomenon.

Systematic reviews of machine-learning-based science have identified data leakage as one of the most common and consequential sources of irreproducible findings, with a recent survey identifying leakage in several hundred published studies across seventeen scientific fields [11]. Reporting frameworks for prediction-model studies, including the TRIPOD+AI statement, now explicitly call for transparent reporting of how such leakage was assessed and excluded [12]. Adversarial validation—training a classifier to distinguish two data partitions in order to quantify and localize distributional differences between them—has been used in applied machine learning to detect and diagnose exactly this kind of leakage and dataset shift, though it has rarely been applied systematically within a clinical diagnostic-classification study [13].

The present study used PCOS classification as a real-world case study to develop and apply a layered acquisition-bias audit framework, designed to separate diagnostic information originating from raw database schema, patterns of measurement acquisition, recorded clinical values, and residual signal after controlling for measurement ascertainment and demographic differences between groups. Rather than aiming to produce a clinically deployable diagnostic model, the objective was methodological: to determine how much of the apparent classification performance in a representative retrospective PCOS cohort was attributable to data-acquisition artifacts rather than to genuine physiological information, and to establish whether the resulting framework could be validated under controlled conditions and generalized to other retrospective machine-learning applications in medicine.

## 2. Methods

### 2.1. Study design and objectives

This retrospective methodological study investigated whether machine-learning models developed to distinguish women with PCOS from controls captured disease-related physiological information or primarily exploited differences in data provenance and clinical data acquisition. The study was not designed to develop or validate a diagnostic tool for clinical deployment. Instead, PCOS classification was used as a real-world case study for evaluating acquisition-related bias in retrospective medical machine learning.

We developed a layered acquisition-bias audit framework that progressively separated information originating from the raw database schema, patterns of test availability, recorded laboratory values, and residual clinical information retained after semantic harmonization and control of measurement coverage. The framework compared models based on raw and harmonized missingness patterns, measured values with and without explicit missingness indicators, and ascertainment-balanced feature sets restricted to variables with comparable availability in PCOS and control participants. Additional analyses addressed age-related cohort differences, the limited number of controls, model calibration, and the stability of conclusions under resampling and controlled semi-synthetic perturbations.

The primary objective was to determine whether PCOS status could be predicted from measurement-availability patterns alone, without access to any clinical or laboratory values. The secondary objectives were to: (i) quantify the contribution of source-specific schema differences by comparing raw and semantically harmonized data representations; (ii) evaluate the incremental predictive contribution of explicit missingness indicators relative to measured values; (iii) assess whether discrimination persisted after restricting the analysis to ascertainment-balanced variables and excluding age; (iv) examine the robustness of model performance to control-sample size, repeated age matching, common age support, bootstrap resampling, and label permutation; and (v) evaluate, under known data-generating conditions, whether the proposed framework could distinguish genuine biological signal from schema-related and workflow-related artifacts.

Together, these analyses were designed to determine whether high apparent classification performance reflected clinically meaningful patient information or properties of how the retrospective dataset had been assembled and documented.

### 2.2. Data source and study population

The study used a retrospective clinical database assembled from records of women evaluated at the Department of Endocrinological Gynecology, Medical University of Silesia in Katowice, Poland, between January 2018 and May 2025. The PCOS cohort comprised women undergoing routine endocrine and gynecological assessment, including clinical examination, hormonal testing, metabolic evaluation, thyroid-function testing, and hematological measurements. Because the data originated from routine clinical practice, the availability and documentation of individual measurements varied across participants.

PCOS was diagnosed according to the 2003 Rotterdam ESHRE/ASRM criteria, requiring the presence of at least two of the following after exclusion of alternative etiologies: oligo- or anovulation, clinical and/or biochemical hyperandrogenism, and polycystic ovarian morphology on ultrasound. Alternative causes of hyperandrogenism or menstrual dysfunction were excluded as part of the diagnostic work-up.

The final integrated dataset contained 1,331 records: 1,286 women with confirmed PCOS and 45 non-PCOS comparison records. The comparison records were retained as the control group for the methodological classification experiments. Because the objective of the study was to audit whether models exploited differences in data provenance and acquisition rather than to construct a clinically deployable diagnostic model, the original imbalance and naturally occurring measurement-availability patterns were preserved in the primary analyses. The resulting PCOS- to-control ratio was approximately 28.6:1.

All records were anonymized before analysis, and direct patient identifiers were excluded from the analytical feature set. The study was conducted in accordance with the Declaration of Helsinki and was approved by the Bioethics Committee of Wroclaw Medical University, Poland (approval No. KB-254/2021). Written informed consent was obtained from the participants in accordance with the approved study procedures; for participants younger than 18 years, consent was additionally obtained from a parent or legal guardian.

### 2.3. Data provenance and schema audit

Before model development, the source database underwent a predefined, outcome-agnostic provenance and schema audit. The original matrix contained 1,331 records and 224 recorded variables spanning demographic, endocrine, metabolic, thyroid, hematological, inflammatory, and derived clinical measures. Because the database had been assembled from routine clinical records, variable availability and documentation structure differed substantially across participants.

Administrative and technical fields were identified using predefined column names and naming patterns related to patient identifiers, document labels, record numbers, hospital admission and discharge information, and source-specific metadata. These variables were excluded from all clinical modeling because they could encode patient identity, record provenance, or healthcare-process information unrelated to physiology. Their identification formed part of the provenance audit but did not constitute performance-based feature selection.

The raw schema was then examined for redundant, duplicated, or synonymous representations of the same clinical concept. Several laboratory parameters were recorded under alternative names, abbreviations, or reporting conventions, including multiple columns for Anti-Müllerian hormone, glucose measurements, gonadotropins, thyroid-stimulating hormone, dehydroepiandrosterone sulfate, testosterone, sex hormone-binding globulin, and hematological indices. Such aliases were retained in the raw-schema representation to quantify source- and schema-related leakage, while their subsequent consolidation was performed separately through the predefined clinical harmonization procedure. The coexistence of alternative columns with different group-specific availability was treated as a potential provenance fingerprint rather than as independent biological information.

For every variable, measurement coverage was defined as the proportion of participants with a non-missing value and was calculated overall and separately for the PCOS and control groups. The audit recorded entirely empty variables, highly sparse variables, variables observed only in one group, and the absolute and signed between-group differences in coverage. Missingness was additionally represented as a participant-by-variable binary matrix to evaluate whether systematic patterns of test availability aligned with diagnostic-group membership. The source dataset showed extensive sparsity, including 13 entirely empty columns and a large proportion of variables with more than 90% missing observations, supporting the need to distinguish information contained in recorded values from information contained in the data-acquisition pattern itself.

### 2.4. Clinical-concept harmonization

To distinguish source-specific schema effects from clinically meaningful information, raw laboratory variables were harmonized using a predefined and version-controlled mapping dictionary. Each dictionary entry linked an original database column to a canonical clinical concept and specified the source unit, target unit, conversion rule, alias priority, and clinical approval status. Harmonization rules were established before model evaluation and were not selected or modified on the basis of classification performance.

Measurements representing the same biological parameter but stored under different names, abbreviations, or reporting conventions were mapped to a common feature identifier. This applied, among others, to alternative representations of Anti-Müllerian hormone, glucose measurements, luteinizing hormone, follicle-stimulating hormone, thyroid-stimulating hormone, dehydroepiandrosterone sulfate, testosterone, sex hormone-binding globulin, and selected hematological variables. The presence of such parallel columns in the source matrix was considered a potential source-provenance fingerprint rather than evidence of distinct biological measurements. The original database contained multiple overlapping representations of clinically equivalent variables, including separate columns for the same hormonal markers.

Where necessary, numerical values were converted to a common target unit using predefined multiplicative and additive transformations. Only mappings with confirmed variable identity and approved unit compatibility were included in the harmonized dataset. Entries with ambiguous biological meaning, incompatible units, or insufficient documentation were excluded rather than combined.

When more than one approved alias was populated for the same participant, values were consolidated according to a prespecified source-priority hierarchy. The highest-priority available measurement was retained. Concurrently recorded aliases were compared using predefined absolute and relative tolerances. Discrepant values exceeding these tolerances were recorded in an alias-conflict report and required an explicit resolution rule. Unresolved conflicts, unapproved mappings, or undefined unit conversions caused the publication pipeline to stop, preventing silent consolidation of potentially non-equivalent measurements.

This process yielded two complementary analytical representations. The raw-schema representation preserved the original clinical column structure after exclusion of administrative variables and was used to quantify discrimination attributable to source-specific naming, column splitting, and documentation patterns. The harmonized-concept representation collapsed approved aliases into standardized clinical variables and was used to evaluate whether group-dependent acquisition patterns and value-based discrimination persisted after removal of schema-level redundancy. Comparing these representations allowed schema leakage to be separated from residual workflow-related missingness and from information contained in the recorded clinical values.

### 2.5. Layered acquisition-bias framework

A layered modeling framework was used to determine how much discriminatory information originated from database structure, measurement availability, recorded clinical values, and residual biological signal after controlling for acquisition differences. The framework was applied separately to the raw-schema and harmonized-concept representations described above, allowing source-specific column structure to be distinguished from persistent differences in clinical test acquisition.

At the first level, models were trained using binary missingness masks only, where each feature indicated whether a measurement was recorded for a given participant. No numerical laboratory values were provided. The raw-schema missingness model captured both source-specific column structure and group-dependent test availability, whereas the harmonized missingness model evaluated whether acquisition-related discrimination persisted after synonymous columns had been consolidated into common clinical concepts. Classification above chance in either setting indicated that diagnostic status was encoded in the pattern of available measurements independently of the measured values themselves.

At the second level, models were trained using the recorded clinical values. Values-only models used median imputation without adding explicit missingness indicators, while combined models received both the imputed values and binary indicators identifying originally missing observations. This comparison was designed to assess whether explicit availability information increased model performance beyond the information retained in the value matrix after imputation. Values-only models were not interpreted as entirely free of missing information, because systematic imputation can indirectly preserve acquisition patterns by replacing unrecorded measurements with repeated constants. Such artifacts may remain highly discriminative even after explicit missingness indicators have been removed.

At the final level, modeling was restricted to an ascertainment-balanced panel derived from the harmonized representation. A feature was eligible when it was recorded in at least 70% of participants in each diagnostic group and the absolute difference in coverage between groups did not exceed 10 percentage points. Eligibility was determined exclusively within the training portion of each validation split. This restriction reduced the possibility that classification performance was driven by tests performed predominantly in one group. Two versions of the ascertainment-balanced analysis were evaluated: one retaining all eligible variables, including age, and a more restrictive version excluding age to reduce the direct influence of demographic differences between the PCOS and control cohorts.

Together, the successive model layers formed a prespecified attenuation sequence from the most acquisition-sensitive representation to the most restrictive clinically comparable representation: raw missingness, harmonized missingness, values without explicit indicators, values combined with missingness indicators, ascertainment-balanced harmonized values, and ascertainment-balanced harmonized values without age. Comparisons between these layers were used to localize the source of apparent classification performance rather than to identify a single optimal diagnostic model.

### 2.6. Model development and internal validation

Two complementary supervised learning algorithms were used throughout the acquisition-bias framework: class-weighted logistic regression and class-weighted random forest. Logistic regression represented a linear and comparatively transparent classifier, whereas random forest was included to capture nonlinear effects and interactions without requiring their prior specification. The use of both algorithms was intended to determine whether the identified acquisition-related signals were model-independent rather than to select a single optimal diagnostic model.

Logistic regression was fitted using the liblinear solver, balanced class weights, a maximum of 150 iterations, and a convergence tolerance of 10 ^3^ . Random forest models comprised 30 trees, with a maximum tree depth of 6, a minimum terminal-node size of 2, and balanced-subsample class weighting. A fixed random seed of 42 was used throughout. Hyperparameters were specified before the final analyses and were not optimized against the reported validation results.

All model specifications were implemented as integrated scikit-learn pipelines. Depending on the analytical representation, the pipeline incorporated fold-specific ascertainment-balanced feature selection, median imputation, optional generation of binary missingness indicators, feature standardization for logistic regression, and classifier fitting. Random forest models were fitted without feature scaling. This structure ensured that all data-dependent transformations were estimated using the training data only.

Internal validation was performed using repeated stratified five-fold cross-validation with three complete repetitions. Stratification preserved the relative proportions of PCOS and control participants within each fold, which was particularly important given the limited number of controls. In each repetition, every participant contributed to a held-out validation fold and received an out-of-fold predicted probability. Fold-level performance estimates were retained, while repeated out-of-fold predictions were aggregated at participant level for selected summary and calibration analyses.

All preprocessing steps were performed independently within each training fold. Medians used for imputation, parameters used for standardization, the set of generated missingness indicators, and eligibility for the ascertainment-balanced panel were therefore derived without access to the corresponding validation observations. For coverage-restricted models, group-specific measurement coverage was recalculated in every training fold, and only variables satisfying the prespecified ascertainment criteria within that fold were passed to the subsequent pipeline stages.

The same cross-validation partitions were reused across competing feature representations and both learning algorithms. Consequently, raw missingness, harmonized missingness, values-only, values-plus-missingness, and ascertainment-balanced models were evaluated on identical training and validation samples. This paired validation design reduced variability attributable to different data splits and allowed differences in performance to be attributed more directly to the analytical representation and preprocessing strategy.

### 2.7. Bias-controlled and robustness analyses

A prespecified set of bias-controlled analyses was conducted to determine whether the conclusions of the layered framework depended on the definition of comparable ascertainment, age differences between groups, or the limited size of the control cohort. All analyses retained the leakage-safe pipeline described above, including fold-local feature eligibility, imputation, scaling, and model fitting.

Five bias-controlled scenarios were evaluated. C1 applied a strict ascertainment criterion, requiring at least 80% measurement coverage in each diagnostic group and an absolute between-group coverage difference not exceeding 5 percentage points. C2 applied the primary 70/70/10 criterion and excluded age from the predictor set. C3 used a relaxed criterion requiring at least 60% coverage in both groups and allowing an absolute coverage difference of up to 15 percentage points. C4 addressed age imbalance through repeated age matching. C5 restricted the analysis to the common observed age support of the PCOS and control groups.

For repeated age matching, each control participant was matched to a PCOS participant using nearest-neighbor matching on age within a maximum caliper of 0.5 years. Matching was repeated 10 times using different random seeds to account for the presence of multiple eligible PCOS comparators. Age was excluded from the subsequent predictor matrix, and the matched cohorts were evaluated using stratified cross-validation. This analysis reduced differences in age distribution without allowing the classifier to use age directly.

In the common-support analysis, the study population was restricted to the overlapping age range observed in both groups. The lower boundary was defined as the greater of the two group-specific minimum ages, and the upper boundary as the smaller of the two group-specific maximum ages. Models were subsequently fitted using the harmonized representation with ascertainment-balanced feature selection and age excluded.

Sensitivity to the limited number of controls was examined through a repeated small-control stress test. Balanced subsets were generated using 20, 30, 40, and 45 controls, with an equal number of PCOS participants sampled for each analysis. Five independently sampled balanced cohorts were generated for every control-group size. Within each sampled cohort, performance was estimated using stratified two-fold cross-validation without repetition. Logistic regression and random forest were evaluated across the principal raw, harmonized, and ascertainment-balanced representations.

The stress test was designed to separate instability arising from the small absolute number of controls from effects of the original class imbalance. For every configuration, the distribution of model performance and the proportion of iterations in which the model remained estimable were recorded. When no feature satisfied the required coverage criteria within a training sample, the corresponding ascertainment-balanced model was designated non-estimable. Such iterations were retained in the audit record and summarized through model-estimability rates rather than being silently discarded or assigned artificial performance values.

Together, the C1–C5 scenarios and the small-control stress test assessed whether the principal findings remained consistent under stricter and more relaxed definitions of balanced measurement availability, exclusion of age, repeated demographic matching, restriction to common demographic support, and reduction of the effective cohort size.

### 2.8. Statistical inference and performance assessment

Model performance was evaluated using complementary measures of discrimination, class-specific precision, threshold-dependent classification, and probabilistic accuracy. The primary performance measure was the area under the receiver operating characteristic curve (ROC-AUC). Because the dataset was strongly imbalanced, average precision was calculated separately with PCOS treated as the positive class and with controls treated as the positive class, together with the corresponding class-prevalence baselines. Threshold-dependent performance was summarized using balanced accuracy, sensitivity, specificity, and the Matthews correlation coefficient. Probabilistic performance was additionally assessed using the Brier score and logarithmic loss. Ordinary accuracy was not used as a principal performance measure because it would be dominated by the majority PCOS class.

Uncertainty in performance estimates was quantified using stratified full-pipeline bootstrap resampling. Within each bootstrap iteration, PCOS and control participants were resampled with replacement within the diagnostic group, and the complete analytical workflow was repeated, including ascertainment-balanced feature selection, imputation, scaling, generation of missingness indicators where applicable, model fitting, and performance estimation. The final executed analysis used 20 full-pipeline bootstrap iterations. For each metric, point estimates were accompanied by percentile-based 95% confidence intervals. The number of requested, successful, failed, and non-estimable iterations was retained for every model configuration.

Differences between model representations were evaluated using paired bootstrap contrasts. Competing models were fitted within the same bootstrap samples so that every contrast represented a within-resample performance difference rather than a comparison between independently generated samples. Prespecified comparisons included values plus missingness versus values only, combined information versus missingness only, raw versus harmonized missingness, harmonized values plus missingness versus harmonized values only, ascertainment-balanced models with versus without age, and the conventional raw model versus the most restrictive ascertainment-balanced model without age. For each contrast, the paired difference distribution was summarized using the median and a percentile-based 95% confidence interval. These contrasts were interpreted as comparisons between complete modeling strategies and not as an additive decomposition of ROC-AUC.

Label-permutation testing was used to determine whether discrimination exceeded that expected under the null hypothesis of no association between the predictors and PCOS status. Diagnostic labels were randomly permuted while the predictor matrix and missingness structure were preserved, after which the complete cross-validation pipeline was rerun. Permutation testing was performed for the principal raw missingness, harmonized missingness, raw values-plus-missingness, and ascertainment-balanced age-excluded configurations. The final executed analysis used 20 permutations for each tested specification.

The empirical permutation P value was calculated as:

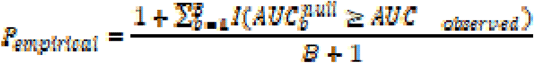

where B denotes the number of successful permutations. Permutation summaries included the observed ROC-AUC, the mean and standard deviation of the null distribution, its upper quantiles, the number of successful permutations, and the empirical P value. With 20 permutations, the smallest attainable empirical P value was 1/21 = 0.0476.

Calibration was evaluated separately using out-of-fold predicted probabilities, calibration plots, the Brier score, expected calibration error, and calibration intercept and slope where estimable. Uncertainty in selected calibration measures was assessed using 200 bootstrap iterations. All performance summaries were derived from held-out predictions only, and apparent training performance was not used for statistical inference.

### 2.9. Semi-synthetic validation

A semi-synthetic experiment was conducted to determine whether the proposed framework could distinguish genuine biological discrimination from artifacts introduced by source-specific schema structure and group-dependent data acquisition. This analysis complemented the retrospective clinical case study by providing a controlled environment in which the mechanisms generating the observed signal were known in advance.

Each simulated dataset contained 600 observations and a set of continuous latent clinical variables generated from prespecified distributions. Three factors were varied in a full factorial design: biological signal strength, workflow-related missingness shift, and source-specific schema shift. Biological signal strength was set to 0, 0.4, or 0.8 and was introduced through class-dependent differences in the latent clinical values. Workflow shift was set to 0, 0.3, or 0.6 and was generated by making the probability that a measurement was recorded dependent on the diagnostic class. Schema shift was set to 0, 0.3, or 0.7 and was introduced by splitting selected latent clinical concepts into alternative source-specific columns whose use differed between groups. Each factorial condition was repeated three times using independent random seeds.

The simulated data were analyzed in two representations. The raw representation retained source-specific aliases as separate columns and therefore preserved both schema-related and workflow-related effects. The harmonized representation collapses aliases derived from the same latent clinical concept into a common feature, reproducing the clinical-concept harmonization strategy used in the empirical analysis. This enabled direct assessment of whether semantic harmonization attenuated discrimination attributable specifically to schema fragmentation while leaving workflow-related and biological signals intact.

For each simulated condition, three input representations were evaluated: missingness indicators only, imputed clinical values without explicit missingness indicators, and imputed values combined with missingness indicators. The semi-synthetic experiment used the logistic-regression pipeline with the same preprocessing logic as the empirical analysis. Each simulated dataset was divided using a stratified 70:30 training–test split, with the split determined by the repetition-specific random seed. Model preprocessing was fitted on the training portion and applied unchanged to the held-out test portion.

The experiment was designed around four prespecified expectations. First, missingness-only models were expected to remain near chance when neither workflow nor schema shift was present. Second, their discrimination was expected to increase as workflow or schema shift increased, even when no biological signal was present. Third, harmonization was expected to reduce the component of performance attributable to source-specific alias splitting while having limited effect on discrimination caused by class-dependent measurement availability. Fourth, value-based models were expected to retain discrimination after harmonization when a genuine biological signal was present.

Performance was summarized across repetitions for every combination of biological, workflow, and schema effects. The principal outputs were ROC-AUC distributions for each input representation before and after harmonization, together with contrasts quantifying the change in discrimination following alias consolidation. The semi-synthetic analysis was interpreted as a mechanism-validation experiment for the audit framework rather than as an attempt to reproduce the exact distribution of the clinical cohort.

### 2.10. Software and reproducibility

All analyses were implemented in Python using a modular computational workflow designed for execution in Google Colaboratory or in an equivalent local environment. Core data processing and statistical procedures used pandas, NumPy, SciPy, scikit-learn, and statsmodels. Matplotlib was used for figure generation, openpyxl for spreadsheet outputs, and PyArrow for storage of harmonized analytical matrices. Required software dependencies were specified in both requirements.txt and environment.yml files.

All principal analytical settings were stored in a version-controlled JSON configuration file. These settings included the random seed, cross-validation design, model parameters, ascertainment thresholds, age-matching criteria, stress-test settings, resampling parameters, simulation settings, and publication-mode status. The final analysis was executed with fast_mode=false and a fixed random seed of 42. The effective configuration used during execution was copied to the output directory, and run metadata recorded the complete configuration, execution mode, and completion status. Technical smoke-test settings and outputs were stored separately and were not used as publication estimates.

Reproducibility safeguards were incorporated throughout the pipeline. All preprocessing and feature-selection operations were performed within the validation procedure, non-estimable model configurations were recorded explicitly, and failed iterations were written to dedicated logs. Automated completion checks verified the presence and basic integrity of the required outputs before the analytical run was classified as complete.

Full implementation details — module structure, automated test coverage, directory layout, and the complete reproducibility manifest (environment, SHA-256 checksums of data, configuration, and outputs) — are provided in the code repository and Supplementary Material rather than in the main text.

## 3. Results

### 3.1. Cohort composition and source-data structure

The final integrated dataset comprised 1,331 records, including 1,286 women with confirmed PCOS and 45 non-PCOS control records. PCOS cases therefore represented 96.6% of the cohort, whereas controls accounted for 3.4%, corresponding to a PCOS-to-control ratio of approximately 28.6:1. The marked imbalance was retained in the primary analyses because it reflected the composition of the retrospective source database rather than an experimentally balanced diagnostic cohort. The cohort composition is summarized in Table 1 and visualized in Figure 1A.

**Figure 1.**
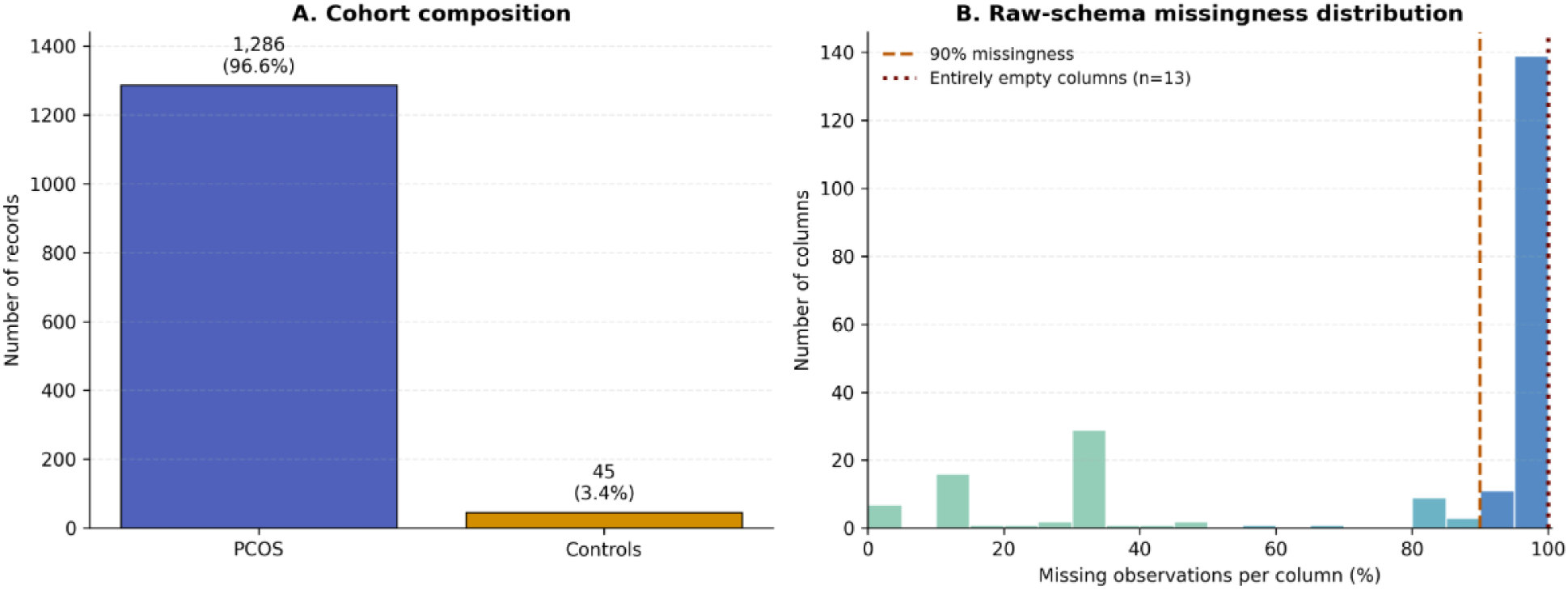
Cohort composition and raw-schema sparsity. (A) Distribution of PCOS and control records in the final integrated dataset. Percentages refer to th total study cohort of 1,331 records. (B) Distribution of the percentage of missing observations across the 224 raw source columns. The dashed vertical line indicates 90% missingness, and the dotted line indicates entirely empty columns.

**Table 1.** Cohort composition and source-data structure.

| Domain | Measure | Value | Notes |
| --- | --- | --- | --- |
| Cohort | Total records | 1,331 |  |
| Cohort | PCOS records | 1,286 |  |
| Cohort | Control records | 45 |  |
| Cohort | PCOS proportion | 96.6% | Percentage of all records |
| Cohort | Control proportion | 3.4% | Percentage of all records |
| Cohort | PCOS-to-control ratio | 28.6 : 1 | PCOS records per control record |
| Source schema | Raw columns | 224 | Includes target and administrative fields |
| Source schema | Numeric candidate features | 182 | After excluding target and administrative fields |
| Source schema | Administrative columns | 2 | Nr KG, subject_id |
| Sparsity | Entirely empty columns | 13 | 100% missing |
| Sparsity | Columns with $\geq 99\%$ missingness | 79 | Across the raw 224-column schema |
| Sparsity | Columns with $\geq 90\%$ missingness | 150 | Across the raw 224-column schema |
| Sparsity | Median column-level missingness | 96.7% | Across the raw 224-column schema |

The original source matrix contained 224 columns. After exclusion of the diagnostic target and two predefined administrative variables (Nr KG and subject_id), 182 numeric clinical features were available for modeling. The retained variables represented a heterogeneous set of demographic, endocrine, metabolic, thyroid, hematological, inflammatory, imaging-related, and derived clinical measurements. The administrative fields were excluded before modeling because they could directly encode patient identity or record provenance.

The raw database was characterized by extensive sparsity. Thirteen of the 224 source columns were entirely empty, 79 columns had at least 99% missing observations, and 150 columns had at least 90% missing observations. Median column-level missingness was 96.7%, indicating that more than half of the raw variables were recorded for fewer than approximately 3.3% of participants. The distribution of missingness across the raw schema is shown in Figure 1B. This pronounced sparsity, together with the substantial imbalance between PCOS and control records, established the conditions under which patterns of test availability could function as strong proxies for diagnostic-group membership.

### 3.2. Clinical-concept harmonization and coverage asymmetry

The predefined harmonization dictionary contained 35 approved raw-to-concept mappings, which were consolidated into 17 canonical clinical concepts. All mapping entries passed the automated clinical and technical validation checks, and no unapproved variables entered the harmonized analytical matrix. The harmonized concepts included age, gonadotropins, androgen-related measurements, thyroid-stimulating hormone, glucose measurements, cortisol, prolactin, and selected hematological indices. The harmonization process and the resulting concept-level coverage are summarized in Table 2.

**Table 2.** Clinical-concept harmonization and group-specific measurement coverage.

| **Panel A. Harmonization summary** |  |  |
| --- | --- | --- |
| Approved raw-to-concept mappings | 35 | All dictionary entries used in the publication pipeline were clinically approved |
| Unapproved mappings | 0 | No unapproved mapping entered the harmonized matrix |
| Canonical clinical concepts generated | 17 | Approved raw aliases were consolidated into standardized concepts |
| Rows with more than one populated alias | 1 | Participant rows in which multiple approved aliases were simultaneously present |
| Conflicts outside tolerance | 1 | Discordant alias values detected during harmonization |
| Unresolved conflicts | 0 | All detected conflicts were resolved according to prespecified rules |
| Harmonization validation status | PASS | Automated validation outcome |
| **Panel B. Coverage asymmetry summary** |  |  |
| Harmonized concepts evaluated | 17 | Concepts included in group-specific coverage analysis |
| Concepts with FDR-adjusted $q < 0.05$ | 16 | Significant differences in measurement availability between PCOS and controls |
| Concepts without significant coverage difference | 1 | Age was the only concept with identical complete coverage |

Only one participant record contained more than one simultaneously populated alias for the same canonical concept. This occurred for fasting glucose, for which the two available source columns contained values of 97 and 89 mg/dL. The discrepancy exceeded the predefined absolute and relative tolerances and was retained in the alias-conflict report. It was resolved using the prespecified source-priority rule, leaving no unresolved alias conflicts and resulting in an overall harmonization validation status of PASS.

Despite removal of source-specific naming and alias fragmentation, substantial group-dependent differences in measurement availability remained. Sixteen of the 17 harmonized concepts showed a statistically significant difference in coverage between the PCOS and control groups after false-discovery-rate correction. Age was the only concept with identical and complete coverage in both groups.

The strongest control-enriched acquisition patterns were observed for platelet count, red-cell distribution width, and mean platelet volume. Each of these measurements was available for all 45 controls but for only 44 of 1,286 PCOS records, corresponding to coverage of 100.0% and 3.4%, respectively, and an absolute difference of 96.6 percentage points. Conversely, 120-minute glucose after oral glucose loading was recorded for 1,149 PCOS participants but for none of the controls, producing coverage of 89.3% and 0%, respectively.

Additional hematological concepts also remained more frequently available in controls. Neutrophil and lymphocyte counts were recorded for 70.1% of PCOS records and 100.0% of controls, while white blood cell count was available for 73.5% and 100.0%, respectively. Differences for the principal hormonal concepts were smaller but remained systematic: PCOS coverage ranged from approximately 90.6% to 91.1%, compared with complete coverage among controls. Fasting glucose was available for 89.5% of PCOS participants and all controls. The complete group-specific coverage pattern is shown in Figure 2.

**Figure 2.**
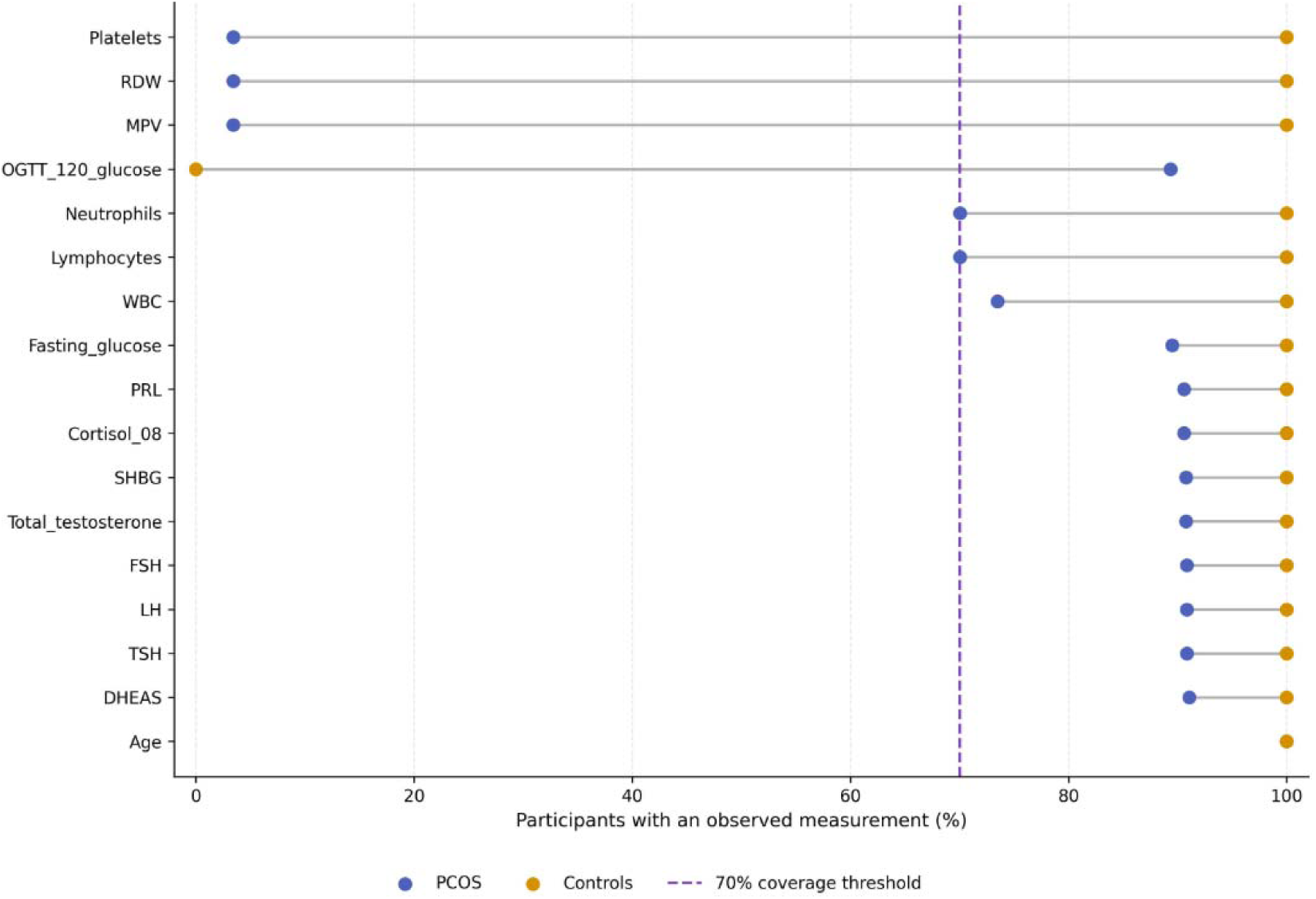
Group-specific measurement coverage after clinical-concept harmonization. Legend: RDW -red cell distribution width; MPV - mean platelet volume; OGTT_120_glucose - glucose after OGTT; WBC - white blood cells; PRL - prolactin; Cortisol_08 - cortisol at 8 a.m.; SHBG - sex hormone-binding globulin; FSH-follicle-stimulating hormone; LH-luteinizing hormone; TSH - thyroid-stimulating hormone; DHEAS-dehydroepiandrosterone sulfate.

### 3.3. Diagnostic status was predictable from missingness alone

Diagnostic-group membership was perfectly discriminated using binary measurement-availability patterns without access to any recorded clinical or laboratory values. Across repeated stratified five-fold cross-validation, both logistic regression and random forest achieved a mean ROC-AUC of 1.000 in all 15 held-out folds for the raw-schema missingness representation. Average precision was 1.000 with either PCOS or controls treated as the positive class, and balanced accuracy, sensitivity, specificity, and the Matthews correlation coefficient were all equal to 1.000 (Table 3).

**Table 3.**
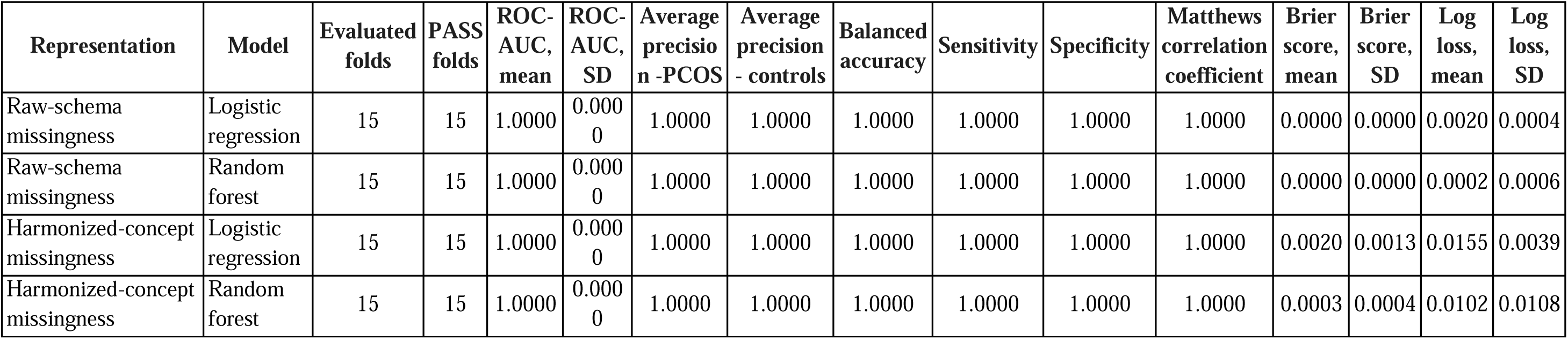
Diagnostic classification performance using missingness patterns alone.

Perfect discrimination was not eliminated by clinical-concept harmonization. Models trained exclusively on the 17 harmonized missingness indicators also achieved a ROC-AUC of 1.000 in every validation fold for both algorithms. Balanced accuracy, sensitivity, specificity, and class-specific average precision likewise remained equal to 1.000. Thus, collapsing source-specific aliases into canonical clinical concepts removed schema fragmentation but did not remove the diagnostic information contained in the pattern of which clinical measurements had been obtained.

The out-of-fold probability distributions showed complete separation between diagnostic groups (Figure 3).

**Figure 3.**
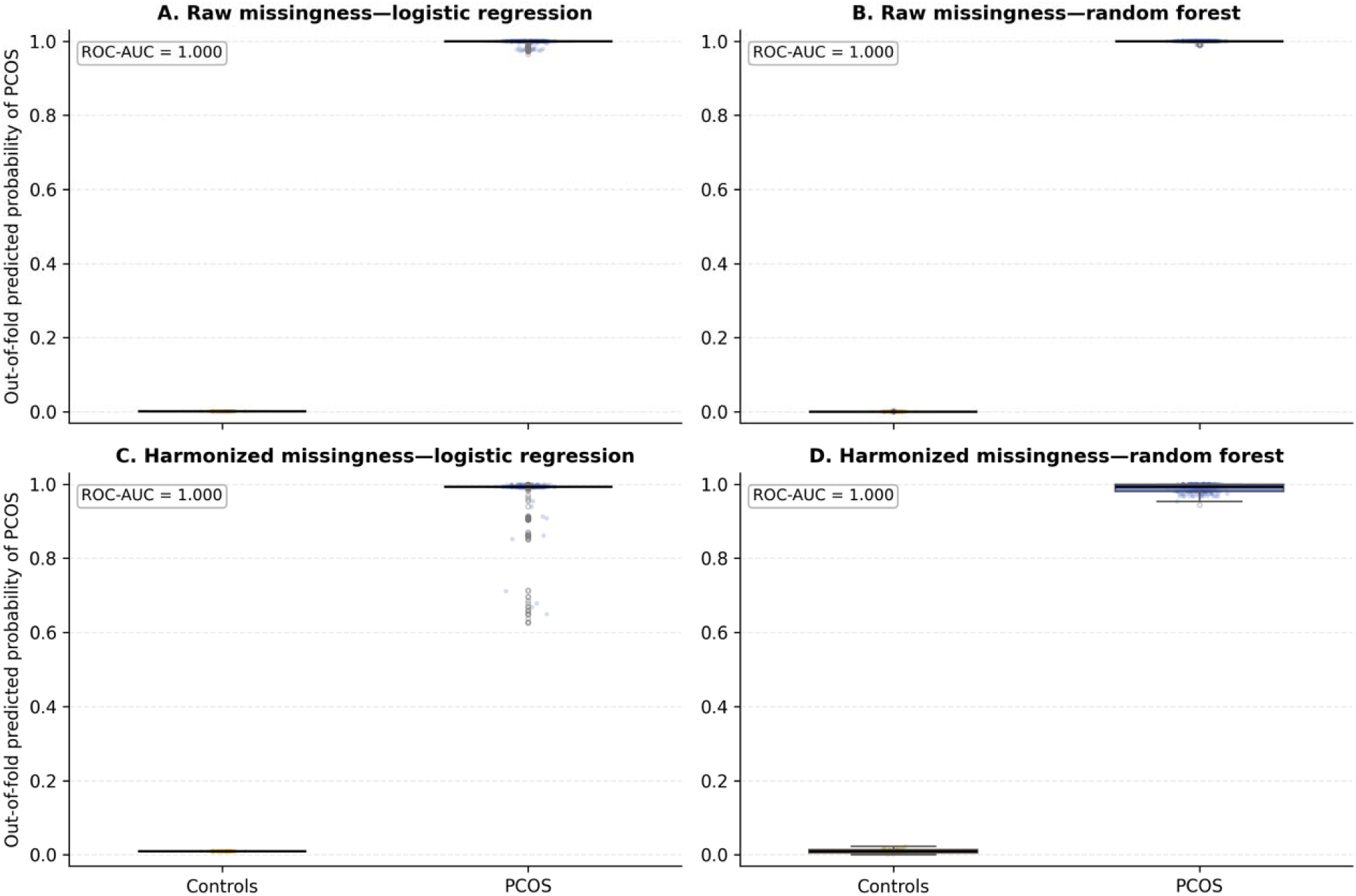
Diagnostic status predicted from missingness patterns alone. Participant-level out-of-fold probabilities of PCOS obtained using binary missingness indicators without access to clinical values. (A) Raw-schema missingness with logistic regression. (B) Raw-schema missingness with random forest. (C) Harmonized-concept missingness with logistic regression. (D) Harmonized-concept missingness with random forest. Probabilities were averaged across the three cross-validation repetitions for each participant. All four configurations achieved a ROC-AUC of 1.000.

For raw-schema missingness, logistic-regression probabilities were concentrated near zero for controls and near one for participants with PCOS. Random forest produced an even more polarized separation, assigning probabilities of approximately zero to controls and one to almost all PCOS records. Following harmonization, separation remained complete, although the logistic-regression predictions for a small subset of PCOS records were less extreme. The lowest participant-level mean probability assigned to a PCOS record was 0.626 for harmonized logistic regression, compared with a maximum of 0.010 among controls

Probabilistic-error measures were also close to zero. For the raw-schema representation, the mean Brier score was 0.000036 for logistic regression and 0.000005 for random forest. The corresponding mean log losses were 0.002047 and 0.000161. After harmonization, mean Brier scores increased to 0.001958 for logistic regression and 0.000299 for random forest, while mean log losses were 0.015499 and 0.010190, respectively. Despite this modest reduction in probability extremity, all observations remained correctly ranked and classified in every validation fold.

Because the models received only binary indicators of whether each measurement was present or absent, these results demonstrate that the acquisition pattern alone contained sufficient information to reproduce PCOS and control status in the source cohort.

### 3.4. Layered model comparisons

Model performance remained close to the ceiling when the raw clinical matrix was used, even after explicit missingness indicators had been removed. In the raw values-only representation, logistic regression achieved a mean ROC-AUC of 0.9995 across the 15 held-out folds, whereas random forest achieved a mean ROC-AUC of 1.000. Adding explicit missingness indicators to the raw values did not improve discrimination: the corresponding mean ROC-AUC values remained 0.9995 for logistic regression and 1.000 for random forest. Balanced accuracy was similarly unchanged, indicating that the information encoded by the raw acquisition structure remained recoverable from the imputed value matrix even without explicit missingness flags (Table 4; Figure 4).

**Figure 4.**
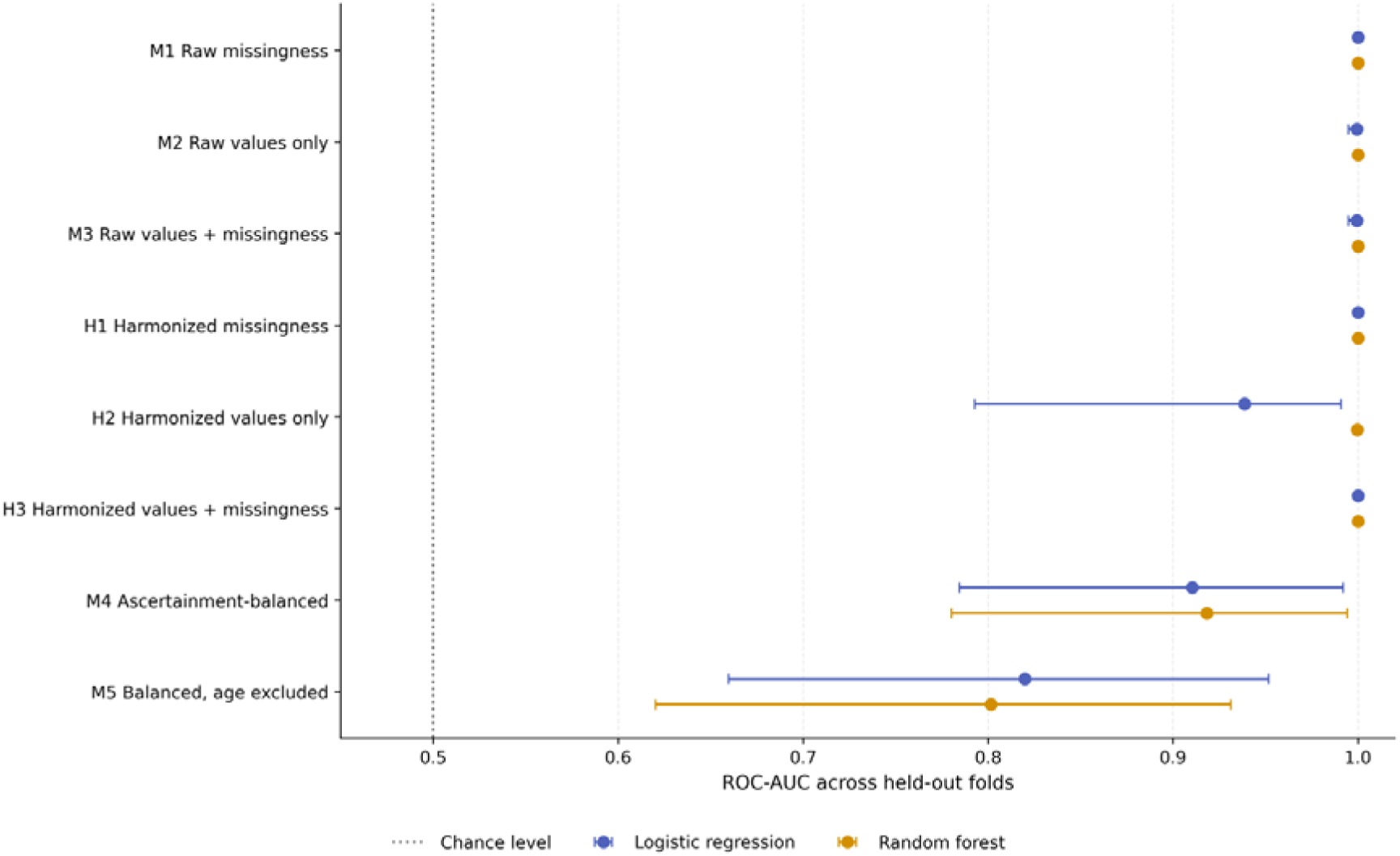
Layered attenuation of apparent classification performance.

**Table 4.** Layered comparison of classification performance across raw, harmonized, and ascertainment-balanced data representations.

| Model layer | Algorithm | Mean ROC-AUC | SD ROC-AUC | 2.5th percentile | 97.5th percentile | Mean AP (PCOS) | Mean AP (controls) | Mean balanced accuracy | Mean sensitivity | Mean specificity | Mean MCC | Mean Brier score | Mean log loss | Folds |
| --- | --- | --- | --- | --- | --- | --- | --- | --- | --- | --- | --- | --- | --- | --- |
| M1 Raw missingness | Logistic regression | 1.0000 | 0.0000 | 1.0000 | 1.0000 | 1.0000 | 1.0000 | 1.0000 | 1.0000 | 1.0000 | 1.0000 | 0.0000 | 0.0020 | 15 |
| M1 Raw missingness | Random forest | 1.0000 | 0.0000 | 1.0000 | 1.0000 | 1.0000 | 1.0000 | 1.0000 | 1.0000 | 1.0000 | 1.0000 | 0.0000 | 0.0002 | 15 |
| M2 Raw values only | Logistic regression | 0.9995 | 0.0020 | 0.9950 | 1.0000 | 1.0000 | 0.9775 | 0.9997 | 0.9995 | 1.0000 | 0.9934 | 0.0005 | 0.0184 | 15 |
| M2 Raw values only | Random forest | 1.0000 | 0.0000 | 1.0000 | 1.0000 | 1.0000 | 1.0000 | 1.0000 | 1.0000 | 1.0000 | 1.0000 | 0.0004 | 0.0081 | 15 |
| M3 Raw values + missingness | Logistic regression | 0.9995 | 0.0020 | 0.9950 | 1.0000 | 1.0000 | 0.9775 | 0.9997 | 0.9995 | 1.0000 | 0.9934 | 0.0005 | 0.0181 | 15 |
| M3 Raw values + missingness | Random forest | 1.0000 | 0.0000 | 1.0000 | 1.0000 | 1.0000 | 1.0000 | 1.0000 | 1.0000 | 1.0000 | 1.0000 | 0.0000 | 0.0003 | 15 |
| H1 Harmonized missingness | Logistic regression | 1.0000 | 0.0000 | 1.0000 | 1.0000 | 1.0000 | 1.0000 | 1.0000 | 1.0000 | 1.0000 | 1.0000 | 0.0020 | 0.0155 | 15 |
| H1 Harmonized missingness | Random forest | 1.0000 | 0.0000 | 1.0000 | 1.0000 | 1.0000 | 1.0000 | 1.0000 | 1.0000 | 1.0000 | 1.0000 | 0.0003 | 0.0102 | 15 |
| H2 Harmonized values only | Logistic regression | 0.9386 | 0.0644 | 0.7927 | 0.9909 | 0.9970 | 0.6007 | 0.8798 | 0.9152 | 0.8444 | 0.4401 | 0.0669 | 0.2575 | 15 |
| H2 Harmonized values only | Random forest | 0.9997 | 0.0009 | 0.9976 | 1.0000 | 1.0000 | 0.9937 | 0.8741 | 1.0000 | 0.7481 | 0.8572 | 0.0060 | 0.0268 | 15 |
| H3 Harmonized values + missingness | Logistic regression | 1.0000 | 0.0000 | 1.0000 | 1.0000 | 1.0000 | 1.0000 | 1.0000 | 1.0000 | 1.0000 | 1.0000 | 0.0000 | 0.0013 | 15 |
| H3 Harmonized values + missingness | Random forest | 1.0000 | 0.0000 | 1.0000 | 1.0000 | 1.0000 | 1.0000 | 1.0000 | 1.0000 | 1.0000 | 1.0000 | 0.0006 | 0.0049 | 15 |
| M4 Ascertainment-balanced | Logistic regression | 0.9105 | 0.0692 | 0.7845 | 0.9919 | 0.9956 | 0.5134 | 0.8716 | 0.8914 | 0.8519 | 0.3963 | 0.0990 | 0.3541 | 15 |
| M4 Ascertainment-balanced | Random forest | 0.9182 | 0.0705 | 0.7801 | 0.9942 | 0.9954 | 0.6498 | 0.7565 | 0.9946 | 0.5185 | 0.6174 | 0.0236 | 0.1351 | 15 |
| M5 Balanced, age excluded | Logistic regression | 0.8199 | 0.0959 | 0.6597 | 0.9516 | 0.9916 | 0.1816 | 0.7552 | 0.7919 | 0.7185 | 0.2210 | 0.1583 | 0.5367 | 15 |
| M5 Balanced, age excluded | Random forest | 0.8016 | 0.1037 | 0.6201 | 0.9313 | 0.9898 | 0.2356 | 0.6129 | 0.9590 | 0.2667 | 0.1889 | 0.0521 | 0.2251 | 15 |

Clinical-concept harmonization produced a more pronounced separation between values-only and missingness-aware models. When logistic regression was trained using harmonized values without missingness indicators, the mean ROC-AUC decreased to 0.9386, with substantial variation across folds (SD 0.0644; 2.5th–97.5th percentile, 0.7927–0.9909). Mean balanced accuracy was 0.8798. The harmonized values-only random forest retained an almost perfect ranking performance, with a mean ROC-AUC of 0.9997, but its mean balanced accuracy was lower at 0.8741. This discrepancy indicated that near-perfect rank discrimination did not necessarily translate into equally strong classification at the prespecified probability threshold.

Reintroducing explicit missingness indicators into the harmonized feature matrix restored perfect performance for both algorithms. Harmonized values-plus-missingness models achieved a mean ROC-AUC, balanced accuracy, sensitivity, and specificity of 1.000 across all validation folds. The contrast between harmonized values-only and harmonized values-plus-missingness was particularly marked for logistic regression, demonstrating that concept-level test availability provided substantial discriminatory information beyond the recorded values themselves.

The strongest attenuation occurred when the analysis was restricted to variables satisfying the prespecified ascertainment-balance criterion. In the ascertainment-balanced representation, mean ROC-AUC decreased to 0.9105 for logistic regression and 0.9182 for random forest. The corresponding mean balanced accuracies were 0.8716 and 0.7565, respectively. Excluding age from this restricted panel reduced performance further: mean ROC-AUC declined to 0.8199 for logistic regression and 0.8016 for random forest, while mean balanced accuracy decreased to 0.7552 and 0.6129.

The progressive change across analytical layers is shown in Figure 4.

Missingness-only and conventional raw-data models remained at or near perfect discrimination, whereas performance declined after semantic harmonization of values, restriction to comparably ascertained variables, and exclusion of age. This attenuation pattern indicated that a substantial component of the apparent classification signal was associated with measurement availability, raw-schema structure, and demographic differences rather than with a stable multivariable physiological profile.

### 3.5. Bias-controlled analyses

Classification performance remained above chance under all bias-controlled scenarios, but its magnitude and threshold-dependent behavior varied substantially across the applied controls (Supplementary Table S1; Figure 5).

**Figure 5.**
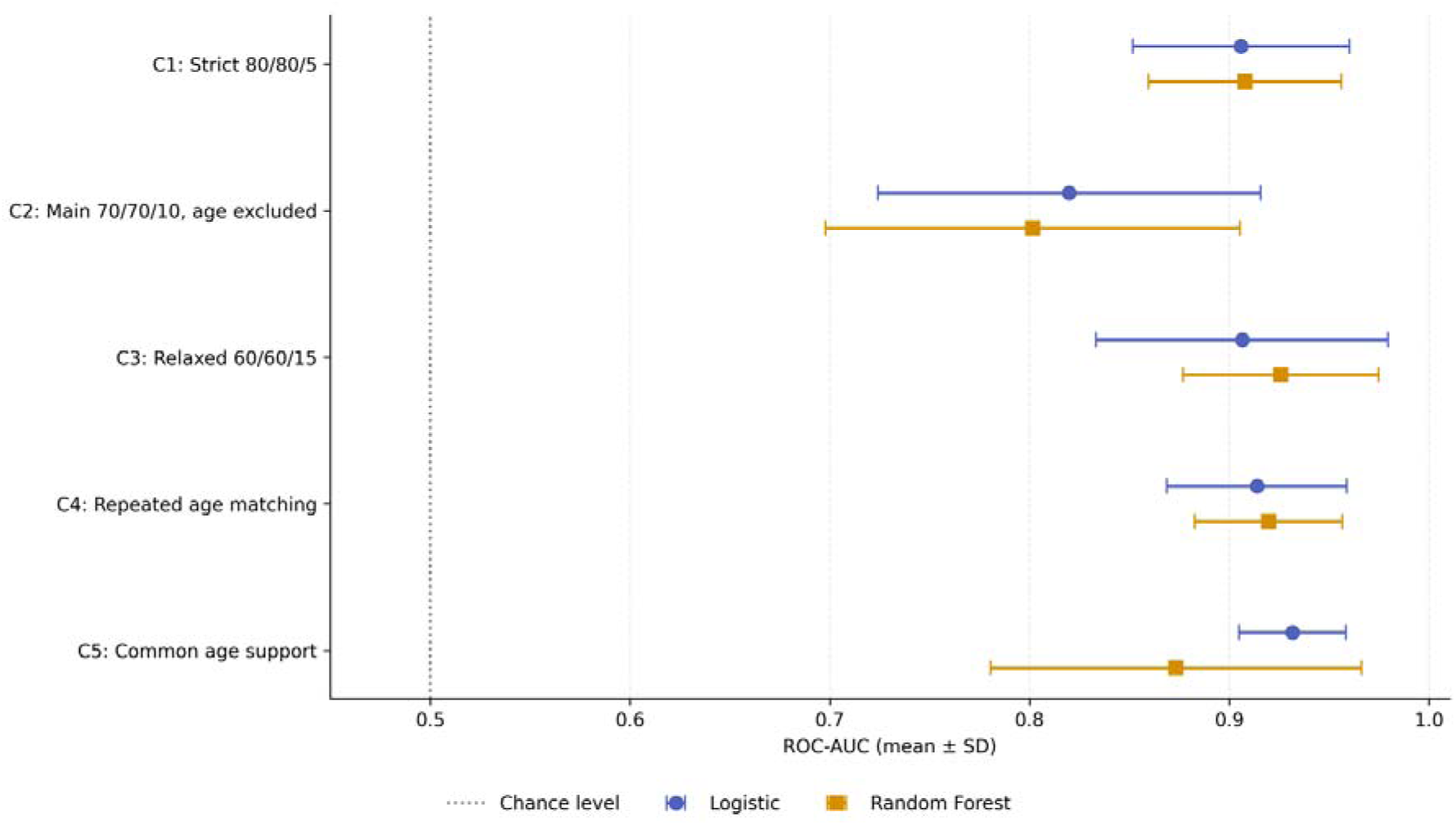
Bias-controlled classification performance.

Under the strict ascertainment criterion requiring at least 80% coverage in both groups and an absolute coverage difference not exceeding 5 percentage points, mean ROC-AUC was 0.906 for logistic regression and 0.908 for random forest. Mean balanced accuracy was 0.828 and 0.837, respectively. The random forest showed higher sensitivity than logistic regression (0.918 versus 0.782) but lower specificity (0.756 versus 0.874), indicating that similar rank discrimination could be accompanied by different classification behavior at the prespecified decision threshold.

Exclusion of age under the primary 70/70/10 ascertainment criterion produced the largest reduction in discrimination among the full-cohort analyses. Mean ROC-AUC declined to 0.820 for logistic regression and 0.802 for random forest. Logistic regression retained a mean balanced accuracy of 0.755, whereas random forest balanced accuracy decreased to 0.613. The latter model remained highly sensitive to PCOS records (mean sensitivity, 0.959) but showed poor control classification (mean specificity, 0.267). These findings indicate that excluding age reduced the residual signal and revealed a pronounced imbalance between case and control classification for the tree-based model.

Relaxing the ascertainment criterion to minimum coverage of 60% in both groups and a maximum coverage difference of 15 percentage points increased mean ROC-AUC to 0.906 for logistic regression and 0.926 for random forest. Logistic regression achieved a mean balanced accuracy of 0.865, with sensitivity and specificity of 0.885 and 0.844, respectively. Random forest achieved near-complete sensitivity (0.994) but lower specificity (0.452), resulting in a mean balanced accuracy of 0.723. Thus, the increase in ROC-AUC under the relaxed criterion did not correspond to balanced classification of both diagnostic groups.

Repeated age matching was performed across 20 independently generated matched cohorts. Each iteration contained 25 control–PCOS pairs. Eighteen iterations were estimable for each algorithm, while two were non-estimable because no feature satisfied the ascertainment criteria within at least one training fold. Across the successful iterations, mean ROC-AUC was 0.914 for logistic regression and 0.920 for random forest. Mean balanced accuracy was 0.841 and 0.833, respectively. Sensitivity and specificity were comparatively balanced in both models, reaching 0.813 and 0.869 for logistic regression and 0.807 and 0.860 for random forest.

The common-age-support analysis restricted the cohort to participants aged 20–25 years, retaining 965 records, including 940 participants with PCOS and 25 controls. Logistic regression achieved a mean ROC-AUC of 0.932 and a mean balanced accuracy of 0.863, with sensitivity of 0.873 and specificity of 0.853. Random forest retained a mean ROC-AUC of 0.873 but showed marked threshold-dependent imbalance: sensitivity was 0.995, whereas specificity was only 0.187, producing a mean balanced accuracy of 0.591.

The distribution of ROC-AUC estimates across the five bias-controlled scenarios is shown in Figure 5.

Discrimination remained above chance after stricter coverage control, age matching, and restriction to common age support. However, the reduction observed after excluding age, together with the low specificity of random forest in several scenarios, showed that residual apparent performance was sensitive to both the available predictor set and the classification algorithm. Consequently, ROC-AUC alone overstated the ability of some models to distinguish both groups reliably.

### 3.6. Robustness to control-sample size

Model behavior was further evaluated in repeatedly sampled balanced cohorts containing 20, 30, 40, or 45 controls and an equal number of PCOS cases. Five independent cohorts were generated for each control-sample size. Performance estimates and model-estimability rates are summarized in Supplementary Table S2 and visualized in Figure 6.

**Figure 6.**
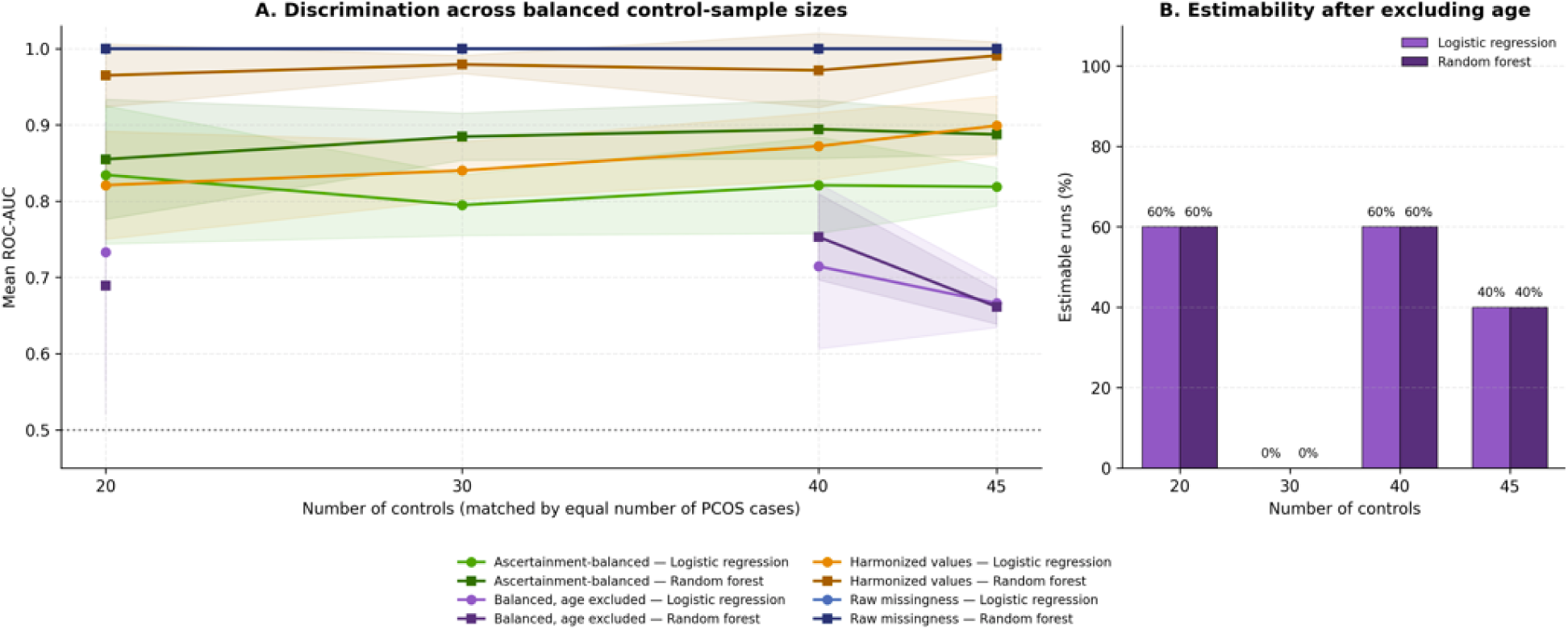
Robustness of model performance to control-sample size. (A) Mean ROC-AUC across five independently generated balanced cohorts containing 20, 30, 40, or 45 controls and an equal number of PCOS cases. Shaded regions represent ±1 standard deviation across estimable iterations. Selected raw, harmonized, and ascertainment-balanced representations are shown for logistic regression and random forest. The horizontal dotted line indicates chance-level discrimination. (B) Proportion of estimable iterations for the ascertainment-balanced, age-excluded representation. Non-estimability occurred when no feature satisfied the predefined coverage criteria in at least one training fold.

Models based on missingness patterns remained highly stable across all evaluated control-sample sizes. Raw-schema missingness achieved a mean ROC-AUC of 1.000 for both logistic regression and random forest at every sample size. Harmonized missingness also remained at or near the performance ceiling. Mean ROC-AUC for harmonized-missingness logistic regression ranged from 0.987 with 30 controls to 1.000 with 20 and 45 controls, while random forest ranged from 0.997 to 1.000. These results indicate that the ability to distinguish the diagnostic groups from measurement-availability patterns was not dependent on the original 28.6:1 class imbalance.

Raw value-based models also retained very high discrimination in the balanced subsets. Random forest trained on raw values achieved a mean ROC-AUC of 1.000 at every evaluated control-sample size. Logistic regression using raw values produced mean ROC-AUC values of 0.960, 0.975, 0.995, and 0.982 for control-sample sizes of 20, 30, 40, and 45, respectively. Adding explicit missingness indicators restored or maintained ceiling-level performance for both algorithms.

Greater variability was observed after clinical-concept harmonization when models were restricted to recorded values without explicit missingness indicators. For logistic regression, mean ROC-AUC increased from 0.821 with 20 controls to 0.899 with 45 controls. The corresponding random-forest estimates remained substantially higher, ranging from 0.965 to 0.991. Nevertheless, harmonized values combined with missingness indicators again produced near-perfect discrimination across all control-sample sizes.

Performance was lower and more variable for the ascertainment-balanced panel. Logistic-regression mean ROC-AUC ranged from 0.795 to 0.835, whereas random-forest mean ROC-AUC ranged from 0.855 to 0.894. Increasing the control-sample size did not produce a consistent monotonic improvement, indicating that variation in fold-specific feature eligibility contributed to performance instability in addition to the absolute number of controls.

The most restrictive representation, which combined ascertainment balancing with age exclusion, was frequently non-estimable because no variable satisfied the coverage criteria in at least one training fold. Both algorithms were estimable in three of five iterations with 20 controls, in none of the five iterations with 30 controls, in three of five iterations with 40 controls, and in two of five iterations with 45 controls (Figure 6B). Among successful iterations, mean ROC-AUC ranged from 0.667 to 0.733 for logistic regression and from 0.661 to 0.753 for random forest. These estimates were substantially lower than those obtained from acquisition-sensitive representations and were accompanied by wide between-iteration variability.

Overall, balancing the number of PCOS and control records did not attenuate the near-perfect performance of missingness-based or conventional raw-data models (Figure 6A). By contrast, models restricted to comparably ascertained clinical information showed lower discrimination, greater variability, and frequent non-estimability after age exclusion. The persistence of ceiling-level missingness performance in balanced samples therefore indicates that the principal signal arose from systematic acquisition differences rather than from the overall class ratio.

### 3.7. Bootstrap inference, paired contrasts, and permutation testing

Full-pipeline bootstrap resampling confirmed that the principal performance pattern was not driven by a small number of individual observations. Missingness-only and missingness-aware models remained at or near the performance ceiling across the bootstrap samples, whereas substantially wider uncertainty was observed for models restricted to ascertainment-balanced clinical information (Supplementary Table S3).

All 20 bootstrap iterations were successfully completed for the raw and harmonized acquisition-sensitive models. Raw-schema missingness achieved a median ROC-AUC of 1.000 with a percentile-based 95% confidence interval of 1.000–1.000 for both logistic regression and random forest. The same result was obtained for harmonized-concept missingness and for harmonized values combined with missingness indicators. Raw values-only logistic regression also remained close to the ceiling, with a median ROC-AUC of 1.000 and a 95% confidence interval of 0.996– 1.000, while the corresponding random-forest model achieved 1.000 across all bootstrap iterations.

Greater uncertainty emerged after explicit acquisition information was reduced. Harmonized values-only logistic regression achieved a median bootstrap ROC-AUC of 0.943 (95% CI, 0.833–0.974), compared with 1.000 (95% CI, 1.000–1.000) after missingness indicators were added. The ascertainment-balanced logistic-regression model achieved a median ROC-AUC of 0.889 (95% CI, 0.839–0.945), while the corresponding random forest achieved 0.926 (95% CI, 0.859–0.963). After age was excluded, one of the 20 bootstrap iterations became non-estimable for each algorithm. Across the 19 successful iterations, median ROC-AUC decreased to 0.816 (95% CI, 0.684–0.894) for logistic regression and 0.809 (95% CI, 0.658–0.885) for random forest.

Paired bootstrap contrasts quantified the contribution of missingness and age within otherwise comparable analytical strategies (Figure 7A). For logistic regression, adding missingness indicators to harmonized values increased ROC-AUC by a mean of 0.071 (95% CI, 0.026– 0.167), increased balanced accuracy by 0.139 (95% CI, 0.067–0.242), and reduced the Brier score by 0.058 (95% CI, 0.042–0.080). For a random forest, the corresponding ROC-AUC contrast was small because the values-only model already achieved near-ceiling rank discrimination: mean difference, 0.001 (95% CI, 0.000 - 0.003). Nevertheless, adding missingness indicators increased balanced accuracy by 0.153 (95% CI, 0.045–0.308) and reduced the Brier score by 0.007 (95% CI, 0.005–0.011).

**Figure 7.**
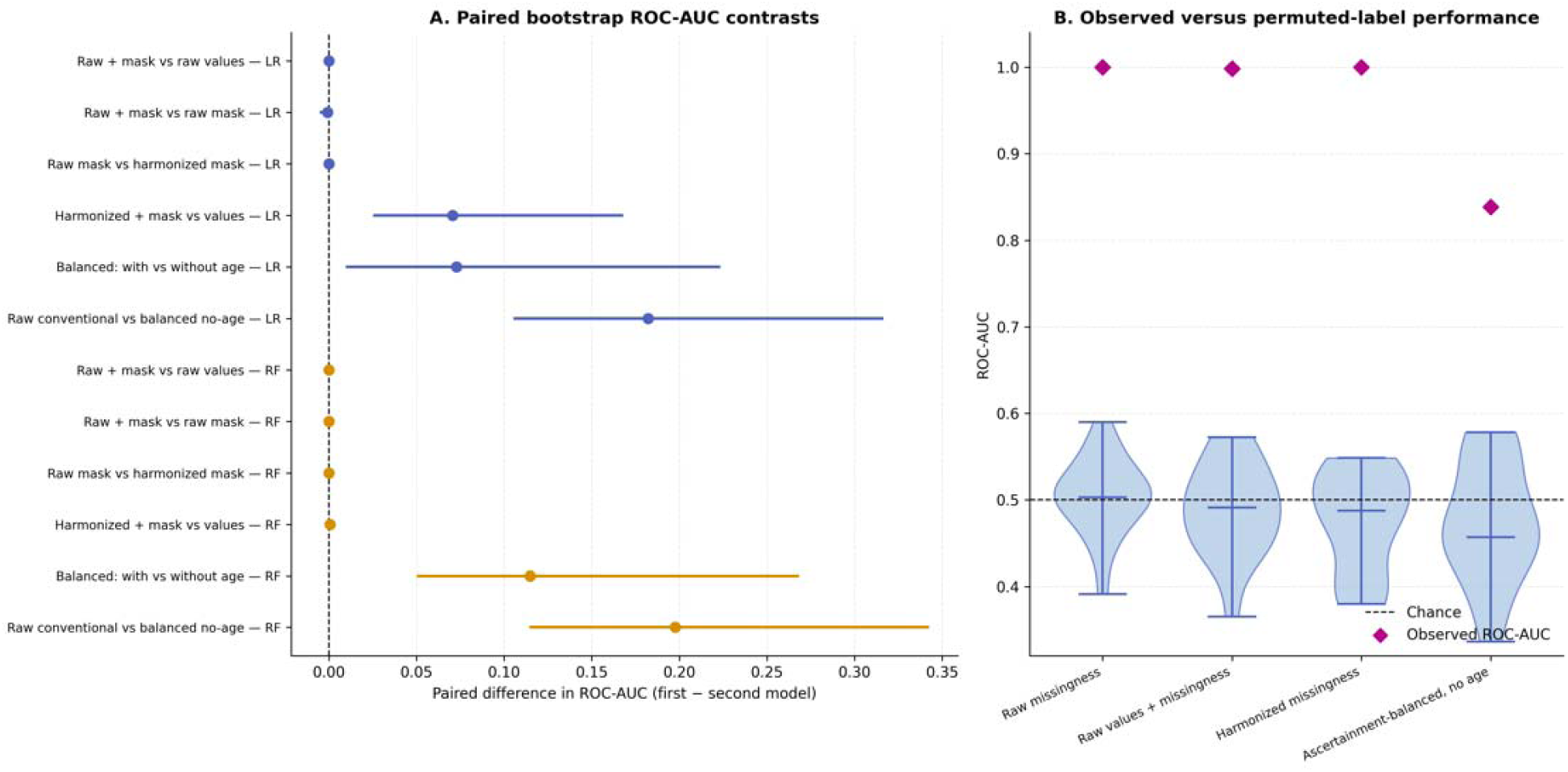
Bootstrap contrasts and label-permutation inference. (A) Paired differences in ROC-AUC between prespecified analytical strategies. Points represent mean within-bootstrap differences, and horizontal lines represent percentile-based 95% confidence intervals. Positive values favor the first model named in each comparison. Blue denotes logistic regression and orange denotes random forest. (B) Null ROC-AUC distributions obtained after permutation of diagnostic labels for four key logistic-regression specifications. Diamonds indicate the observed ROC-AUC, and the horizontal dashed line denotes chance-level discrimination.

Excluding age from the ascertainment-balanced panel reduced ROC-AUC for both algorithms. The paired difference between models with and without age was 0.073 for logistic regression (95% CI, 0.011–0.223) and 0.115 for random forest (95% CI, 0.051–0.268). The corresponding logistic-regression difference in balanced accuracy was 0.101 (95% CI, 0.022–0.218). For random forest, the balanced-accuracy interval included zero despite a positive mean difference of 0.110 (95% CI, −0.039 to 0.228), reflecting instability in threshold-dependent classification.

The largest contrasts were observed between the conventional raw values-plus-missingness representation and the ascertainment-balanced, age-excluded model. Mean ROC-AUC was higher by 0.182 for logistic regression (95% CI, 0.106–0.316) and by 0.198 for random forest (95% CI, 0.115–0.342). Balanced accuracy was higher by 0.257 (95% CI, 0.161–0.367) and 0.393 (95% CI, 0.290–0.515), respectively. In contrast, adding explicit missingness indicators to the raw value matrix produced no measurable change in ROC-AUC or balanced accuracy for either algorithm, indicating that the raw imputed matrix already preserved the acquisition-related signal. Raw and harmonized missingness-only models also had identical ROC-AUC and balanced accuracy.

Label-permutation testing produced null ROC-AUC distributions centered close to 0.5, while the observed values remained substantially higher (Figure 7B). For raw missingness, the observed ROC-AUC was 1.000, compared with a null mean of 0.504, a null standard deviation of 0.045, and a null 95th percentile of 0.559. The empirical PPP value was 0.0476 based on 20 successful permutations.

The raw values-plus-missingness model achieved an observed ROC-AUC of 0.998, compared with a null mean of 0.488 and a null 95th percentile of 0.563. Harmonized missingness achieved an observed ROC-AUC of 1.000, compared with a null mean of 0.475 and a null 95th percentile of 0.548. Both analyses yielded empirical P=0.0476P=0.0476P=0.0476. For the ascertainment-balanced, age-excluded model, the observed ROC-AUC was 0.839, compared with a null mean of 0.475 and a null 95th percentile of 0.578. Nineteen permutations were successful for this specification, resulting in an empirical PPP value of 0.050.

Together, the bootstrap and permutation analyses confirmed that the observed discrimination exceeded that expected after random label assignment. However, the paired contrasts showed that a substantial proportion of the apparent performance was lost after explicit control of measurement availability and age. The strongest and most stable results were therefore obtained from representations that retained information about the diagnostic acquisition process.

### 3.8. Calibration of acquisition-sensitive models

Calibration differed substantially across analytical representations and was strongly influenced by whether the models retained information about measurement availability. Acquisition-sensitive models generally produced very low Brier scores and expected calibration errors, whereas calibration deteriorated after restricting the analysis to harmonized values and comparably ascertained clinical variables (Supplementary Table S4; Figure 8).

**Figure 8.**
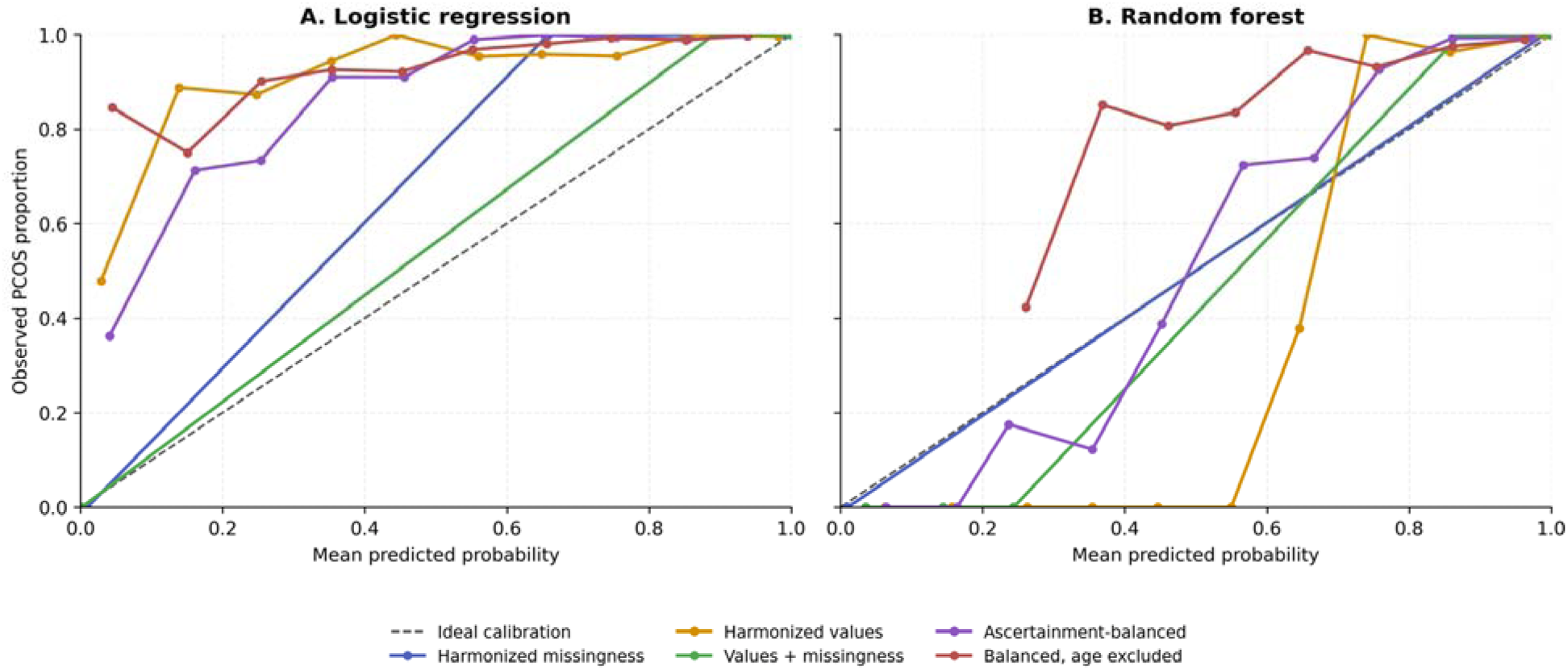
Calibration across acquisition-sensitive and bias-controlled models. Reliability curves comparing mean predicted probabilities with the observed proportion of PCOS for (A) logistic regression and (B) random forest. Curves are shown for harmonized missingness, harmonized values only, harmonized values combined with missingness indicators, the ascertainment-balanced panel, and the ascertainment-balanced panel with age excluded. The dashed diagonal line represents ideal calibration. Calibration estimates were derived from participant-level out-of-fold probabilities, and observed proportions were summarized within adaptive probability bins.

Models based directly on missingness patterns showed near-zero probabilistic error. For harmonized missingness, logistic regression achieved a Brier score of 0.0019 (bootstrap 95% CI, 0.0014–0.0027) and an expected calibration error of 0.0136 (95% CI, 0.0118–0.0160). The corresponding random-forest model achieved a Brier score of 0.00018 (95% CI, 0.00017– 0.00020) and an expected calibration error of 0.0093 (95% CI, 0.0089–0.0099). Similarly low errors were observed for raw-schema missingness, for which the Brier scores were 0.000036 for logistic regression and 0.000002 for random forest.

Adding explicit missingness indicators to harmonized values produced the lowest calibration-error estimates among the harmonized representations. The harmonized values-plus-missingness logistic-regression model achieved a Brier score of 0.000045 (95% CI, 0.000020–0.000075) and an expected calibration error of 0.00115 (95% CI, 0.00081–0.00154). For a random forest, the corresponding values were 0.00051 (95% CI, 0.00032–0.00075) and 0.00095 (95% CI, 0.00009– 0.00222), respectively. The apparent improvement after adding missingness indicators paralleled the restoration of perfect discrimination reported in the preceding analyses.

In contrast, calibration deteriorated when logistic regression was trained using harmonized clinical values without explicit missingness indicators. The Brier score increased to 0.0645 (95% CI, 0.0555–0.0740), and the expected calibration error increased to 0.1077 (95% CI, 0.0948– 0.1177). Its Brier skill score was negative (−0.974), indicating that its probability estimates performed worse than a prevalence-based reference prediction under this metric. The calibration slope was 0.577 (95% CI, 0.439–0.766), consistent with predictions that were too extreme relative to the observed outcomes. The harmonized values-only random forest showed a lower Brier score of 0.0057 (95% CI, 0.0040–0.0078) and an expected calibration error of 0.0084 (95% CI, 0.0044–0.0124), although its recalibration-parameter estimates were unstable.

Restricting the predictors to the ascertainment-balanced panel further exposed calibration limitations. The ascertainment-balanced logistic-regression model had a Brier score of 0.0961 (95% CI, 0.0879–0.1035), an expected calibration error of 0.2270 (95% CI, 0.2140–0.2367), and a negative Brier skill score of −1.941. Its calibration slope was close to one at 0.970 (95% CI, 0.660–1.351), but the calibration intercept was elevated at 3.063 (95% CI, 2.798–3.465), indicating systematic underestimation of the probability of PCOS. The corresponding random forest showed lower error, with a Brier score of 0.0220 (95% CI, 0.0175–0.0262) and an expected calibration error of 0.0408 (95% CI, 0.0353–0.0481).

The poorest calibration was observed after both ascertainment balancing and age exclusion. Logistic regression achieved a Brier score of 0.1541 (95% CI, 0.1420–0.1636), an expected calibration error of 0.2952 (95% CI, 0.2804–0.3063), and a Brier skill score of −3.717. Its calibration slope was 0.442 (95% CI, 0.274–0.734), with an intercept of 3.203 (95% CI, 2.930– 3.493). The corresponding random forest performed better but remained imperfectly calibrated, with a Brier score of 0.0485 (95% CI, 0.0421–0.0547), an expected calibration error of 0.0943 (95% CI, 0.0844–0.1048), and a negative Brier skill score of −0.486.

Reliability curves demonstrated the same pattern (Figure 8). Acquisition-sensitive models concentrated predictions near the extremes and showed very small absolute probability errors because their predicted classes were almost completely separated. By contrast, values-only and bias-controlled models generated predictions across a broader probability range and showed larger deviations from the ideal calibration line. The largest deviations were observed for the ascertainment-balanced, age-excluded models.

The extremely high calibration slopes and intercepts estimated for several acquisition-sensitive models should be interpreted cautiously. These parameters were estimated under complete or near-complete separation, where logistic recalibration coefficients can become numerically extreme and unstable. Accordingly, their low Brier scores and expected calibration errors reflected highly accurate probabilities within this specific source cohort but did not establish transportable calibration for an independent clinical population.

### 3.9. Semi-synthetic validation

The semi-synthetic experiment reproduced the principal mechanisms investigated in the clinical dataset under controlled conditions. A total of 81 simulated datasets were generated across the full factorial combination of three biological-signal levels, three workflow-shift levels, and three schema-shift levels, with three independent repetitions per condition. Each dataset contained 600 observations and was evaluated in raw and harmonized feature spaces using missingness-only, values-only, and values-plus-missingness representations. All 486 planned model evaluations were successfully completed (Table 9).

**Table 9**. Semi-synthetic validation of biological, workflow, and schema effects on classification performance.

When biological, workflow, and schema effects were all absent, discrimination remained close to chance. In the raw feature space, mean ROC-AUC was 0.478 for missingness-only models, 0.532 for values-only models, and 0.506 for values-plus-missingness models. The corresponding estimates after concept harmonization were 0.477, 0.531, and 0.506, respectively. This indicated that the simulation did not generate substantial classification performance in the absence of a prespecified signal.

Schema shift alone created strong apparent discrimination in acquisition-sensitive representations, even when no biological or workflow effect was present. At the maximum schema-shift level of 0.7, the raw missingness-only model achieved a mean ROC-AUC of 1.000, compared with 0.478 under no schema shift. Raw values-plus-missingness likewise increased from a mean ROC-AUC of 0.506 to 0.999. By contrast, the harmonized missingness-only model remained close to chance at 0.477, and harmonized values-plus-missingness remained at 0.506. Thus, consolidation of source-specific aliases almost completely removed discrimination arising solely from schema fragmentation (Figure 9A and 9D).

**Figure 9.**
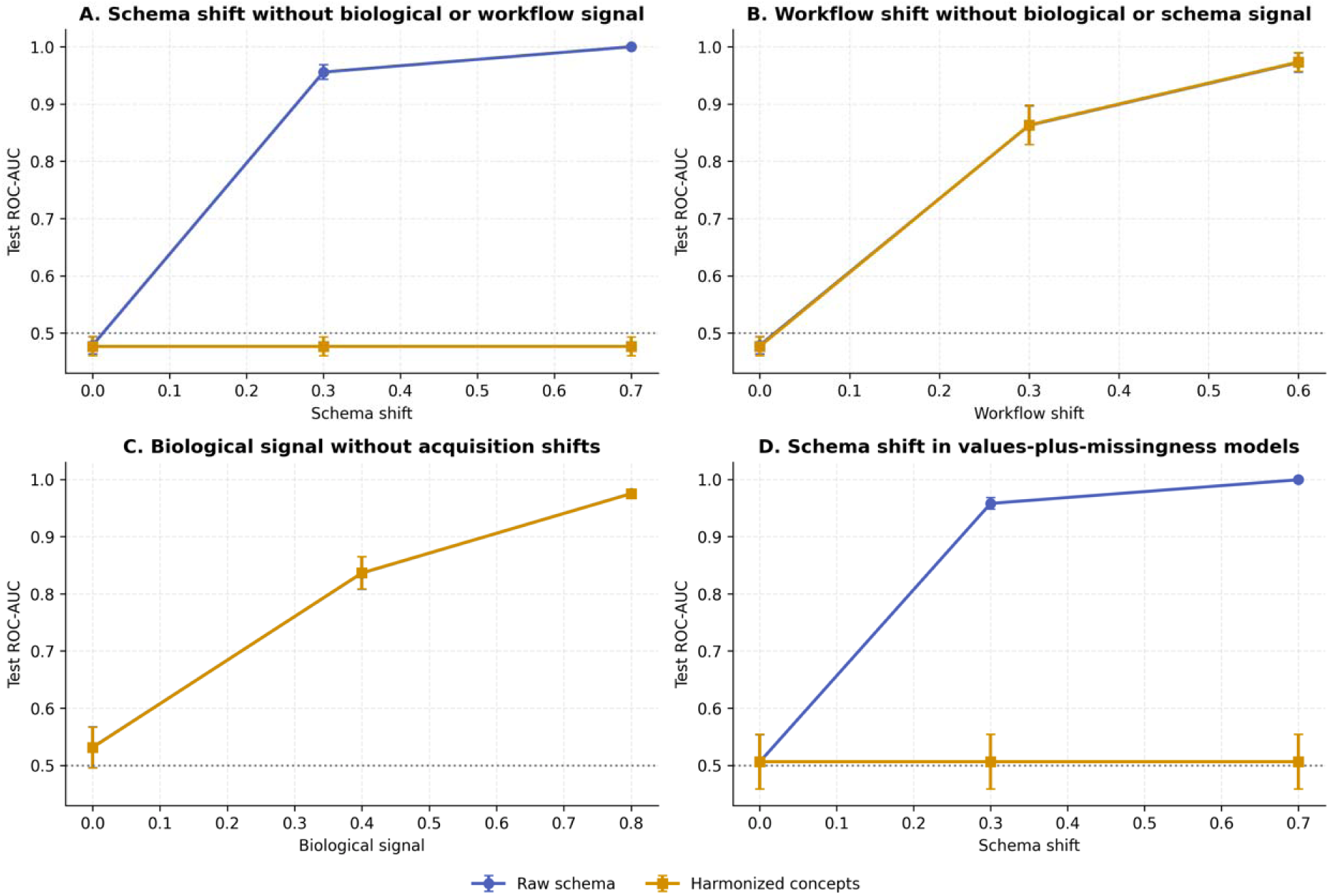
Semi-synthetic validation of biological and acquisition-related signals. Mean held-out ROC-AUC across three independent repetitions; error bars indicate ±1 standard deviation. (A) Effect of source-specific schema shift on missingness-only models in the absence of biological and workflow signals. (B) Effect of workflow-related missingness shift on missingness-only models in the absence of biological and schema signals. (C) Effect of biological-signal strength on values-only models in the absence of acquisition shifts. (D) Effect of schema shift on values-plus-missingness models in the absence of biological and workflow signals. Blue lines represent the raw source schema, and orange lines represent harmonized clinical concepts. The horizontal dotted line denotes chance-level discrimination.

Harmonization did not remove discrimination caused by class-dependent measurement availability. In the absence of biological and schema effects, increasing workflow shift from 0 to 0.3 and 0.6 increased the mean missingness-only ROC-AUC from 0.478 to 0.863 and 0.972 in the raw feature space. Nearly identical values were obtained after harmonization: 0.477, 0.863, and 0.973, respectively (Figure 9B). Values-plus-missingness models showed the same general pattern. These results confirmed that harmonization addressed alias-related schema artifacts but could not eliminate information encoded by the decision to obtain or omit a measurement.

In contrast, values-only models responded primarily to the prespecified biological signal. When workflow and schema shifts were both absent, increasing biological-signal strength from 0 to 0.4 and 0.8 increased mean ROC-AUC from 0.532 to 0.839 and 0.975 in the raw representation. Harmonized values-only models produced nearly identical estimates of 0.531, 0.839, and 0.975 (Figure 9C). This confirmed that consolidation of aliases preserved discrimination derived from genuine class-dependent differences in latent clinical values.

The distinction between biological and acquisition-related information persisted when multiple mechanisms were combined. Under the strongest joint condition—biological signal 0.8, workflow shift 0.6, and schema shift 0.7—the raw missingness-only and values-plus-missingness models both achieved mean ROC-AUC values of approximately 1.000. After harmonization, missingness-only performance remained high at 0.973 because the workflow-related acquisition shift persisted, while values-only performance remained 0.940 because the biological signal was preserved. The harmonized values-plus-missingness model achieved a mean ROC-AUC of 0.995, reflecting the combined contribution of biological values and workflow-dependent availability.

Across the complete factorial experiment, biological-signal strength had no effect on missingness-only performance, whereas workflow and schema shifts strongly increased discrimination in the raw missingness representation. After harmonization, the effect of schema shift disappeared, but the effect of workflow shift remained unchanged. Conversely, values-only performance increased with biological-signal strength and was largely insensitive to workflow or schema shifts. These results matched the prespecified simulation expectations and provided mechanism-level validation of the empirical audit framework.

### 3.10. Summary of principal findings

The acquisition-bias audit produced five convergent findings.

First, the source cohort was characterized by marked class imbalance and extensive raw-schema sparsity. PCOS records represented 96.6% of the cohort, while more than half of the 224 source variables were missing in at least 96.7% of participants.

Second, clinical-concept harmonization successfully removed redundant aliases and source-specific naming differences but did not eliminate group-dependent acquisition patterns. Sixteen of the 17 harmonized concepts showed significant differences in measurement coverage between PCOS and control records, with several hematological measurements predominantly available in controls and 120-minute glucose predominantly available in PCOS records (Table 2; Figure 2).

Third, diagnostic status was perfectly predictable from missingness alone. Both logistic regression and random forest achieved ROC-AUC values of 1.000 using either raw-schema or harmonized-concept missingness indicators.

Fourth, model performance decreased progressively as acquisition-sensitive information was removed. Conventional raw-data models remained at or near the performance ceiling, whereas harmonized values-only models showed lower and more variable performance. Restriction to comparably ascertained variables reduced discrimination further, and exclusion of age produced the lowest full-cohort estimates. The same attenuation pattern was observed in the bootstrap contrasts and remained evident in balanced control-sample stress tests, repeated age matching, and common-age-support analyses (Table 4); Figures 4–7).

Fifth, the semi-synthetic experiment reproduced the mechanisms observed in the clinical data. Schema shift generated high apparent discrimination in raw missingness-aware representations and was removed by harmonization. Workflow-related missingness remained predictive after harmonization, whereas values-only models responded primarily to the simulated biological signal.

Overall, the results showed that near-perfect classification in the source cohort was largely dependent on diagnostic workflow, measurement availability, schema structure, and age. After these sources of information were progressively constrained, discrimination decreased, uncertainty increased, calibration deteriorated, and some models became non-estimable.

## Discussion

This audit of a real-world PCOS cohort found that near-perfect classification performance was, to a large extent, an artifact of how the underlying data had been acquired rather than evidence of a stable, transportable biological signal. Diagnostic status could be recovered with perfect accuracy from the pattern of which measurements were present or absent, before any laboratory value was examined, and this result was not attributable to duplicated or inconsistently named source columns: it persisted, essentially unchanged, after clinical-concept harmonization collapsed 35 raw aliases into 17 canonical concepts. The signal was therefore embedded not in the schema itself but in the underlying clinical measurement workflow — the systematic differences in which tests were ordered for women referred with a working diagnosis of PCOS compared with women assessed for other reasons. As acquisition-sensitive information was progressively removed — first by restricting the analysis to comparably ascertained variables, then by excluding age — apparent discrimination fell from ceiling-level to a mean ROC-AUC of approximately 0.80–0.82, calibration deteriorated markedly, and a subset of bias-controlled and small-control configurations became non-estimable. This attenuation pattern, replicated across two learning algorithms and confirmed under bootstrap resampling, label permutation, and a semi-synthetic experiment with known ground truth, indicates that the ceiling-level performance obtained from raw or lightly processed representations of this dataset substantially overstated the strength of any genuine, clinically transportable classification signal.

### Relation to the wider PCOS machine-learning literature

These findings are difficult to reconcile with the PCOS machine-learning literature at face value. A recent systematic review catalogued numerous studies reporting classification accuracies at or above 95%, most trained and validated within a single, often small and imbalanced, dataset [2]; comparable performance has been reported using widely reused public PCOS datasets [3]. Larger, prospectively assembled cohorts that used electronic-health-record data linked to explicit diagnostic criteria have reported considerably more modest discrimination, in the region of 0.80–0.85 [14]. This gap mirrors the attenuation observed here between acquisition-sensitive and ascertainment-balanced representations of the same cohort, and it raises the possibility that at least part of the very high performance reported elsewhere in the PCOS literature reflects analogous, unaudited acquisition artifacts rather than a stronger underlying biological signal. Because the present dataset, cohort composition, and imbalance are broadly typical of retrospective single-center PCOS studies used for machine-learning research, this concern is unlikely to be idiosyncratic to the present sample.

### A general problem for retrospective clinical machine learning

The mechanism identified here is not specific to PCOS. It belongs to a well-described family of failures in which models trained on retrospective clinical data exploit incidental correlates of the data-generating process rather than the clinical construct of interest — commonly described as shortcut learning [4]. Convolutional networks trained to detect pneumonia or COVID-19 on chest radiographs have been shown to instead recognize the hospital system, department, or scanner that produced an image, because these acquisition-related cues were correlated with disease prevalence in the training data [5,6]; the present results demonstrate an analogous mechanism in structured, tabular clinical data, where the “acquisition fingerprint” is not a pixel-level artifact but the pattern of which laboratory tests were ordered. Because informative missingness of this kind reflects genuine clinical decision-making, it is not intrinsically a modeling error: when a model is intended to operate within the same workflow that generated its training data, missingness indicators can legitimately improve predictive performance [9,10]. The problem arises specifically when a retrospective cohort is used, as here, to make claims about an underlying disease signal rather than to reproduce the behavior of a specific clinical pathway - a distinction that is rarely made explicit in the reporting of PCOS or other disease-classification studies, and one that the present framework was designed to make auditable.

The semi-synthetic experiment reinforces this interpretation and clarifies why semantic harmonization, though necessary, is not sufficient. Harmonization removed discrimination attributable specifically to source-specific alias splitting - the schema-level mechanism - but left workflow-related missingness fully intact, exactly as the empirical clinical analysis showed. This distinction matters for practice: many published pipelines treat “cleaning” of duplicated or inconsistently named variables as sufficient due diligence against data leakage, but the present results indicate that a dataset can pass such cleaning entirely and still encode diagnostic-group membership through the pattern of test ordering alone. Audits that stop at schema harmonization will therefore systematically under-detect this category of leakage.

### Calibration as an additional diagnostic signal

Calibration analysis provided a further, largely underused, diagnostic. Acquisition-sensitive models produced near-zero Brier scores and expected calibration errors, not because their probability estimates were well-calibrated in any transportable sense, but because complete or near-complete separation between groups makes any threshold-independent probabilistic metric appear excellent by construction. Once ascertainment-balanced or age-excluded representations were used, calibration deteriorated sharply and several Brier skill scores became strongly negative, revealing that the ostensibly best-performing models were, in fact, the least trustworthy from a probabilistic standpoint. This observation suggests that unusually good calibration metrics in a retrospective diagnostic-classification study, particularly alongside near-perfect discrimination, should themselves be treated as a signal warranting an acquisition-bias audit rather than as unambiguous evidence of model quality.

### A general framework for auditing acquisition bias in medical machine learning

Beyond the specific PCOS findings, the principal contribution of this work is methodological: a layered audit strategy - comparing missingness-only, value-only, value-plus-missingness, and ascertainment-balanced representations across both raw and harmonized schemas, validated against bootstrap, permutation, and semi-synthetic benchmarks - that is applicable to any retrospective disease-classification task built from routinely collected clinical data. This complements, rather than duplicates, existing leakage taxonomies and reporting frameworks. Kapoor and Narayanan’s taxonomy of leakage in machine-learning-based science identifies “illegitimate features” and non-representative test distributions as recurring failure modes but does not provide a worked procedure for isolating acquisition-workflow leakage from schema leakage in tabular clinical data [11]. The TRIPOD+AI statement calls for transparent reporting of how such risks were assessed but likewise does not specify a concrete analytical pathway [12]. Adversarial validation - training a classifier to distinguish two partitions of a dataset in order to localize distributional differences - has been used productively to detect training-test drift in industrial machine-learning systems [13], and the framework presented here can be understood as a structured, clinically interpretable extension of that idea, decomposed into interpretable layers rather than a single black-box discriminator. We suggest that comparable layered audits — feasible with the standard classifiers and cross-validation procedures used throughout this study - could be adopted as a routine step before reporting diagnostic-classification performance from retrospective medical datasets, particularly when class imbalance, single-center provenance, and heterogeneous test ordering are all present, as they frequently are in real-world clinical machine-learning research.

## Limitations

Several limitations qualify these conclusions. First, the analysis was conducted at a single center, and the control group was small (n=45); although the principal missingness-based findings were stable across repeated small-control resampling and balanced subsets, this cannot substitute for genuine external validation at an independent site. Second, this was a retrospective, cross-sectional dataset without temporal or geographic separation between training and evaluation; the audit framework identifies acquisition-related information within this dataset but cannot, by itself, establish how a model would perform on a truly external cohort. Third, the study was explicitly designed as a methodological audit rather than the development of a clinically deployable diagnostic tool, and no claim is made about the clinical utility of any of the evaluated models. Fourth, the semi-synthetic validation, while reproducing the qualitative mechanisms observed in the clinical data, used simplified latent-variable structures and cannot capture the full complexity of real clinical documentation practice. Finally, the small control group, characteristic of retrospective single-center PCOS databases, limits the precision of some bias-controlled and stress-test estimates, as reflected in the wide confidence intervals and non-estimable iterations reported in the corresponding analyses.

## Conclusions

Near-perfect classification performance in this retrospective PCOS cohort was substantially attributable to diagnostic workflow, schema structure, and measurement-acquisition patterns rather than to a stable and transportable biological signal, and semantic harmonization of the underlying database removed only the schema-related component of this artifact. These findings argue for routine, layered acquisition-bias auditing - extending beyond simple missing-data cleaning - as a standard component of retrospective machine-learning studies in medicine, and the framework developed here provides a concrete, generalizable template for conducting such audits in other disease areas.

## Data Availability

The clinical dataset analyzed during the current study is not publicly available because it contains sensitive patient information and institutional ethical restrictions prohibit unrestricted public release. De-identified data may be made available from the corresponding author upon reasonable request and subject to approval by the appropriate institutional and ethical authorities.

## Code Availability

The complete analytical workflow developed for this study, including Python scripts for data harmonization, preprocessing, missing-data handling, dimensionality reduction, clustering, stability assessment, cross-space agreement analysis, biological validation, sensitivity analyses, and figure generation, is openly available in the following GitHub repository: https://github.com/OlaSt1803/ml_prediction_of_PCOS_or_lab_order

The repository also contains documentation describing the computational workflow, software dependencies, and instructions required to reproduce all analyses presented in this manuscript.

## Ethics Approval

This study was conducted in accordance with the Declaration of Helsinki and was approved by the Bioethics Committee of Wroclaw Medical University (approval no. 254/2021).

## Consent to Participate

Written informed consent was obtained from all participants prior to inclusion in the study.

## Consent for Publication

All authors have reviewed and approved the final version of the manuscript and consent to its publication.

## Author Contributions

Kornelia Kataryńczuk: Code preparation and participated in the first and second review Aleksandra Stachowiak: Code preparation and participated in the first and second review

Natalia Piórkowska: Conceptualization, methodology, software, formal analysis, investigation, visualization, writing – original draft, writing – review and editing.

Alan Ostromęcki: Conceptualization, methodology, visualization, writing – original draft, writing – review and editing.

Grzegorz Franik: Data curation.

Anna Bizoń: Writing – review and editing, supervision. All authors read and approved the final manuscript.

## Data Availability

https://github.com/OlaSt1803/ml_prediction_of_PCOS_or_lab_order

## Acknowledgements

The authors thank all study participants and the clinical staff involved in patient recruitment, data collection, and laboratory diagnostics.

## Supplementary Materials

**Supplementary Table S1.** Bias controlled analyses.

**Supplementary Table S2.** Control sample size robustness.

**Supplementary Table S3.** Bootstrap contrasts and permutation testing.

**Supplementary Table S4.** Calibration of acquisition sensitive models.

